# Immunogenicity of COVID-19 vaccines in patients with haematological malignancy: A systematic review and meta-analysis

**DOI:** 10.1101/2021.11.06.21265967

**Authors:** Joanne S.K. Teh, Julien Coussement, Zoe C. F. Neoh, Tim Spelman, Smaro Lazarakis, Monica A. Slavin, Benjamin W. Teh

## Abstract

The objectives of this study were to assess the immunogenicity and safety of COVID-19 vaccines in patients with haematological malignancy. A systematic review and meta-analysis of clinical studies of immune responses to COVID-19 vaccination stratified by underlying malignancy and published from 1 January 2021 to 31 August 2021 was conducted using MEDLINE, EMBASE and CENTRAL. Primary outcome was the rate of seropositivity following 2 doses of COVID-19 vaccine with rates of seropositivity following 1 dose, rates of positive neutralising antibody (nAb), cellular responses and adverse events as secondary outcomes. Rates were pooled from single arm studies while rates of seropositivity were compared against the rate in healthy controls for comparator studies using a random effects model and expressed as a pooled odds ratio with 95% confidence intervals.

Forty-four studies (16 mixed group, 28 disease specific) with 7064 patients were included in the analysis (2331 following first dose, 4733 following second dose). Overall seropositivity rates were 61-67% following 2 doses and 37-51% following 1 dose of COVID-19 vaccine. The lowest seropositivity rate was 51% in CLL patients and was highest in patients with acute leukaemia (93%). Following 1 dose, nAb and cellular response rates were 18-63% and 33-86% respectively. Active treatment, ongoing or recent treatment with targeted and CD-20 monoclonal antibody therapies within 12 months was associated with poor COVID-19 vaccine immune responses. New approaches to prevention are urgently required to reduce COVID-19 infection morbidity and mortality in high-risk patient groups that respond poorly to COVID-19 vaccination.

## Introduction

The COVID-19 pandemic caused by the SARS-CoV-2 virus has resulted in global mortality of over 3 million deaths. SARS-CoV-2 is likely to remain an endemic viral pathogen, with new variants continuously emerging^1^. There is a significant burden of morbidity and mortality from COVID-19 infection in haematology patients. Over 80% of patients require hospitalisation and up to 50% present with severe disease ^2–5^. Approximately 15% require intensive care unit admission and mortality rates are high at 30-40% depending on underlying disease^2–7^. Poor control of infection due to immune compromise lead to emergence of new variants which further complicate management^8^.

Vaccination is an effective public health measure to reduce risk of infection and severe complications from COVID-19^9^. However, patients with haematological malignancies were excluded from the pivotal trials which preceeded regulatory approvals of the currently recommended COVID-19 vaccines ^10–13^. Understanding the impact of COVID-19 vaccines on cancer patients is critical as social distancing restrictions are being eased in countries with relatively high rates of vaccination coverage^14^. Assessing immune response post-vaccination is the basis of many vaccination studies in patients with haematological malignancy and is often utilised as a surrogate marker of vaccine efficacy^15–17^. These studies are the basis for vaccination recommendations contained within international guidelines^18,19^. Therefore, this study was conducted to systematically review available data on the humoral and cellular immune responses to COVID-19 vaccination in patients with haematological malignancy to build the evidence base for its utility.

## Objectives

The main objective of this systematic review was to assess the immunogenicity (i.e., vaccine-induced immune response) of COVID-19 vaccines in patients with haematological malignancy, stratified by underlying disease type. The secondary objective was to assess the safety of COVID-19 vaccines in the same patient groups.

## Methods

This systematic review was preregistered with PROSPERO (CRD42021276851), and conducted in line with Preferred Reporting Items for Systematic Reviews and Meta-Analyses (PRISMA) guidelines^20^.

### Types of studies and participants

Studies eligible for inclusion into the systematic review were those that investigated the vaccine-induced immunity in patients with haematological malignancy who received at least one dose of COVID-19 vaccine. Randomised controlled trials (RCTs), quasi-RCTs and observational studies were eligible for inclusion. All types of observational studies were included (including prospective and retrospective studies, and studies with or without a control group). This systematic review evaluated all studies that reported on at least one of the following haematology patient groups:

1. All patients with haematological malignancy
2. Patients with multiple myeloma (MM)
3. Patients with chronic lymphocytic leukaemia (CLL)
4. Patients with lymphoma; Hodgkins (HL) and non-Hodgkins lymphoma (NHL)
5. Patients with acute myeloid or lymphocytic leukaemia (AML, ALL)
6. Patients undergoing or who have completed haematopoietic stem cell transplantation (HCT) and chimeric antigen T-cell receptor (CAR-T) therapy
7. Patients with myeloproliferative neoplasm (MPN) and chronic myeloid leukaemia (CML)

### Types of intervention and evaluation

The type of intervention was the use of one or two doses of COVID-19 vaccine (of any type). Immune response to COVID-19 vaccination included humoral and cellular immune responses. For the purpose of this systematic review, humoral response consists of seropositivity which was defined by the SARS-CoV-2 spike/receptor binding domain specific IgG level above the threshold of detection for the assay utilised in each study. Rate of positive neutralising antibody response (nAb) was defined by SARS-CoV-2 specific nAb level above the threshold established by the dedicated neutralisation assay utilised in each study. A positive cellular response was defined by the appropriate increase in frequency of SARS-CoV-2 specific CD4+/CD8+ T cells following vaccination according to assays utilised. Where available, the immune response for each malignant patient group was compared to a control group. If there was no comparator group, the immune response for each haematology patient group was described and summarised.

### Outcome measures

The primary outcome was rates of seropositivity following two doses of COVID-19 vaccine stratified by disease groups while secondary outcome measures were the rates of seropositivity following one dose of vaccine, rates of positive nAb response following one and two doses of COVID-19 vaccine, rates of positive cellular response following one and two doses of COVID-19 vaccine and rates of systemic and/or local adverse events (whichever rate was higher) following two doses of COVID-19 vaccine.

### Search strategy Electronic searching

Literature searches were conducted by an experienced research librarian (SL) using the following databases to identify relevant articles: Ovid Medline, EMBASE and Cochrane CENTRAL from 1 January 2020 to 31 August 2021 and for articles in English only. A combination of subjects and keyword terms encompassing haematologic cancers, COVID-19 and vaccines and all their associated terms was used for the search. The terms used included: haematological neoplasms, leukemia, lymphoma, multiple myeloma, myeloproliferative disorders, myelodysplastic-myeloproliferative diseases, stem cell transplantation, bone marrow transplantation, chimeric antigen receptor therapy, COVID-19, SARS-CoV-2, vaccines, vaccination, immunise, immunisation, BNT162b2, CHAdOx1, AZD 1222, mRNA-1273, Ad26, Ad5, NVX-CoV2373. All word variations were searched and Medical subject headings were exploded. The detailed search strategy is summarised in Supplementary Appendix.

### Selection of studies

Studies were excluded from the systematic review if they did not measure or report immunogenicity after COVID-19 vaccination, if insufficient details were reported for the haematology patient groups in mixed group studies, if there were less than 10 patients including case reports, if data was exclusive to paediatric patients less than 18 years of age or animal studies. Studies that were not peer reviewed and/or published such as pre-prints, abstracts and government reports were excluded. Review articles and other publications without original data such as expert opinions, editorials and consensus statements were also excluded.

Study eligibility was assessed by two independent reviewers (BT, JT), and Covidence systematic review software (Veritas Health Innovation, Melbourne, Australia) was used to screen titles, abstracts, and full texts. Irrelevant reports were discarded, and the full texts of the other reports were accessed. Disagreements between the two main reviewers with respect to study eligibility were resolved by discussion, or via consultation with a third author.

### Data extraction

For studies fulfilling inclusion criteria, data were extracted manually and independently by authors (JT, JC, BT) using a predefined data extraction form. Extracted data elements included study design (enrolment, follow up period, randomisation/allocation, laboratory analysis, predefined outcomes, adjusted analysis, funding source), participant information (inclusion criteria, number of participants, characteristics, disease, treatment), intervention (vaccine type, dose, schedule, comparator group) and outcomes (definitions, timing, adverse events).

### Assessment of risk of bias

Two review authors (ZN, MS) independently assessed the risk of bias of each cohort study using the Newcastle-Ottawa Scale^21^. Each included study was assessed based on three domains: (1) the selection of the study groups, (2) the comparability of the study groups, and (3) the ascertainment of outcome of interest. The rating system proposed by Sharmin et al^22^ was adopted with a good quality study scoring 3 or 4 stars in the selection domain, 1 or 2 stars in comparability domain, and 2 or 3 stars in the outcome domain. For single arm studies, a good quality study would score 3 stars in selection domain, and 2 or 3 stars in outcome domain.

### Measures of treatment effect

For each study, the number of patients achieving seropositivity and the total number of patients receiving vaccination was extracted and expressed as a proportion. For the meta-analysis, rate of response following Ad26 vaccine was analysed at the same time point and therefore summarised as part of a two-vaccine dose response. For each haematology disease group, the proportions from each study were pooled and expressed as an overall proportion (response rate) with 95% confidence intervals (95% CI) utilising inverse variance/generalised mixed modelling.

For each study with a control group, the number of patients with seropositivity in each group and the total number of patients receiving vaccination were extracted, expressed as a proportion with 95% CI and compared against each other. A meta-analysis was performed if there were two or more studies identified with a similar population and sufficient data for the outcomes of interest. The rate of seropositivity as a dichotomous outcome of interest across studies was summarised using a random effects model and expressed as a pooled odds ratio with 95% CI. Analysis was performed using metaprop function on R 4.1.1 (R foundation for Statistical Computing, Vienna).

### Assessment of heterogeneity

Clinical and methodological heterogeneity between included studies was assessed by comparing the key patient factors and study factors (vaccine type, type of assay utilised, threshold for positive response, duration of follow up). Statistical heterogeneity was assessed by using the I^2^ statistic and Chi^2^ with a *p* value ≤ 0.05 considered significant for the presence of heterogeneity.

### Management of missing data

If available, reason(s) for missing data were outlined. Due to time limitations, further information was not sought from the original author/corresponding author(s).

### Subgroup analysis

Pre-planned subgroup analysis was performed to evaluate impact of active treatment (versus no active treatment), anti-CD-20 therapy in the last 12 months including current CD-20 therapy (versus anti-CD-20 therapy 12 or more months ago), targeted therapy (defined as Bruton’s tyrosine kinase inhibitor [BTKi], BCL-2 antagonist venetoclax) in last 12 months (versus no targeted therapy), timing of vaccination in relation to HCT (within 12 months versus longer than 12 months) and response by types of vaccine (BNT162b2 vs others including mRNA-1273 [Spikevax, Moderna], ChAdOx1 [Vaxzevria, AstraZeneca] or Ad26 [Janssen]). For subgroup analysis only studies containing sufficient level of detail for the outcome of interest (e.g. therapy < 12 months) were included. For analysis by vaccine type, only studies reporting use and immune response of multiple vaccines types were included.

## Results

### Search results

A search of electronic databases yielded 520 results and following exclusion of duplicate results, 411 abstracts were reviewed. Forty-eight studies were identified for full text review and subsequently 44 studies fulfilled inclusion criteria and underwent data extraction (Figure 1). Overall there were 44 studies of COVID-19 vaccination in patients with haematological malignancy. Sixteen studies^23–38^ involved mixed group of patients with various underlying haematological malignancies and 28 studies focused on specific malignancies and treatments; 8 myeloma studies^39–46^, 6 CLL studies^47–52^, 4 lymphoma studies^53–56^, 4 studies of HCT^57,58^ and CAR-T^59,60^ and 6 MPN studies^61–66^. A total of 7064 patients were included in the analysis (2331 following first dose and 4733 following second dose). Characteristics of patients, primary and secondary outcomes in included studies are summarised in Table 1.

**Figure 1:**
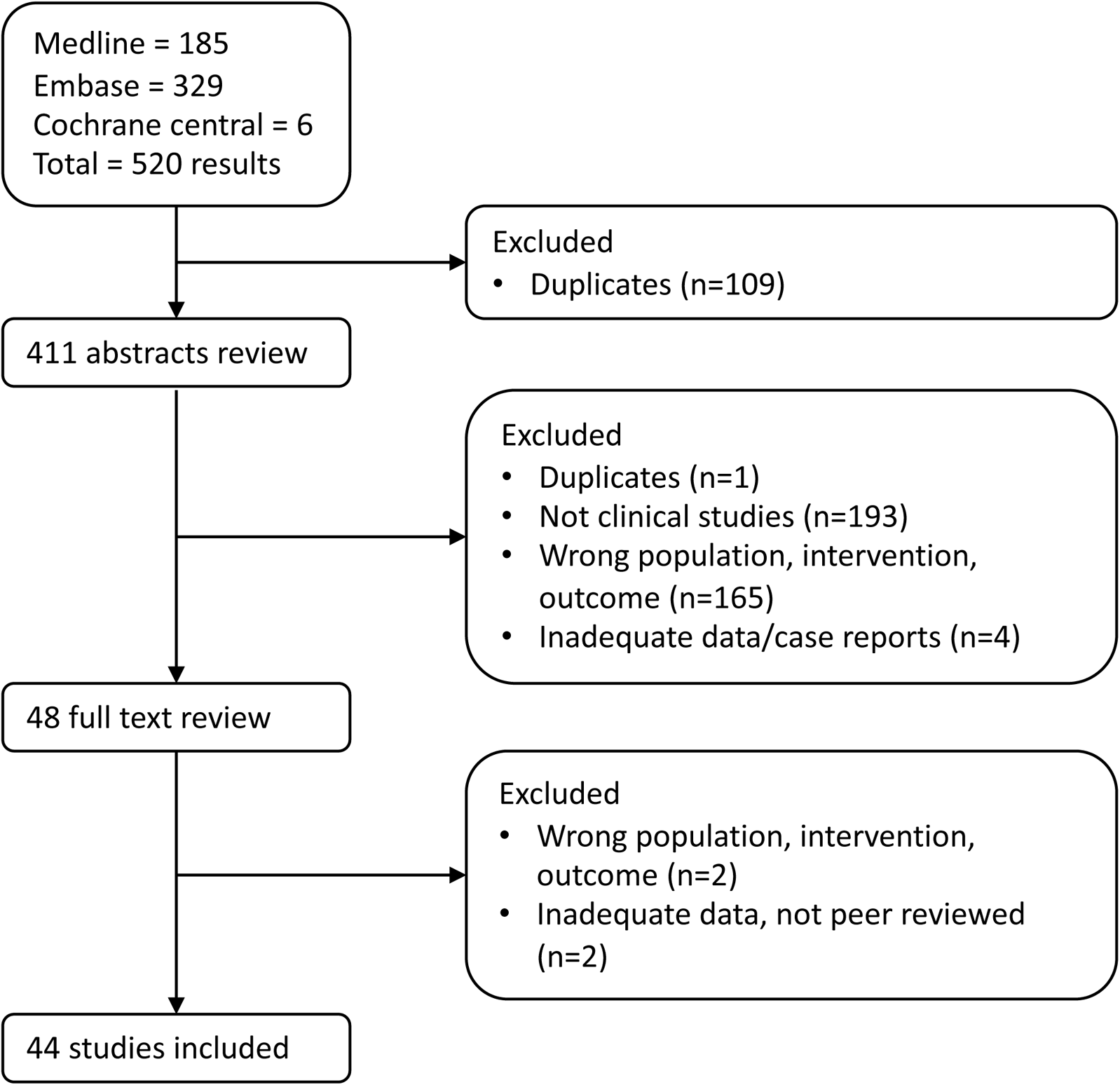
Flow diagram of studies identified, screened and included.

**Table 1:**
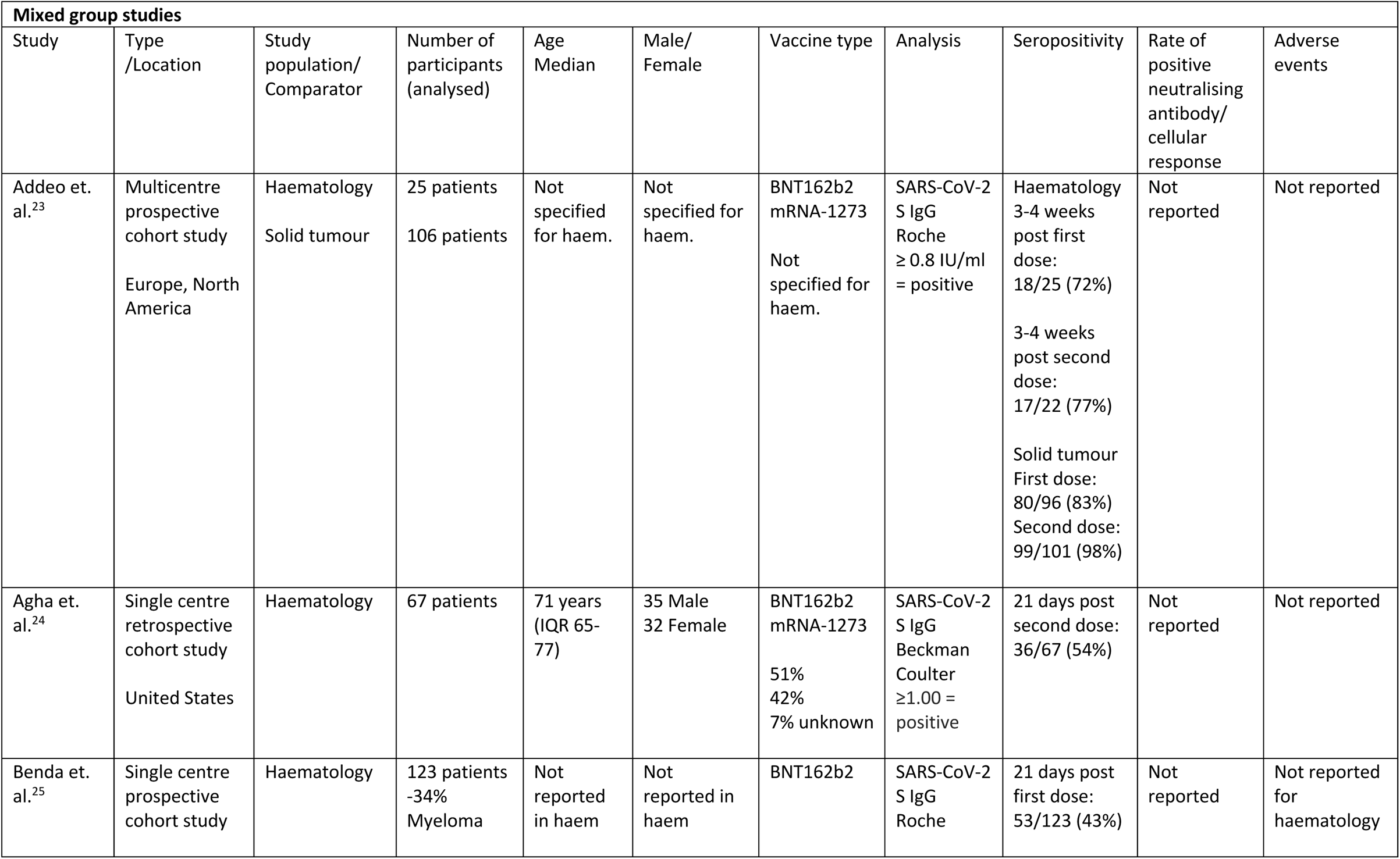
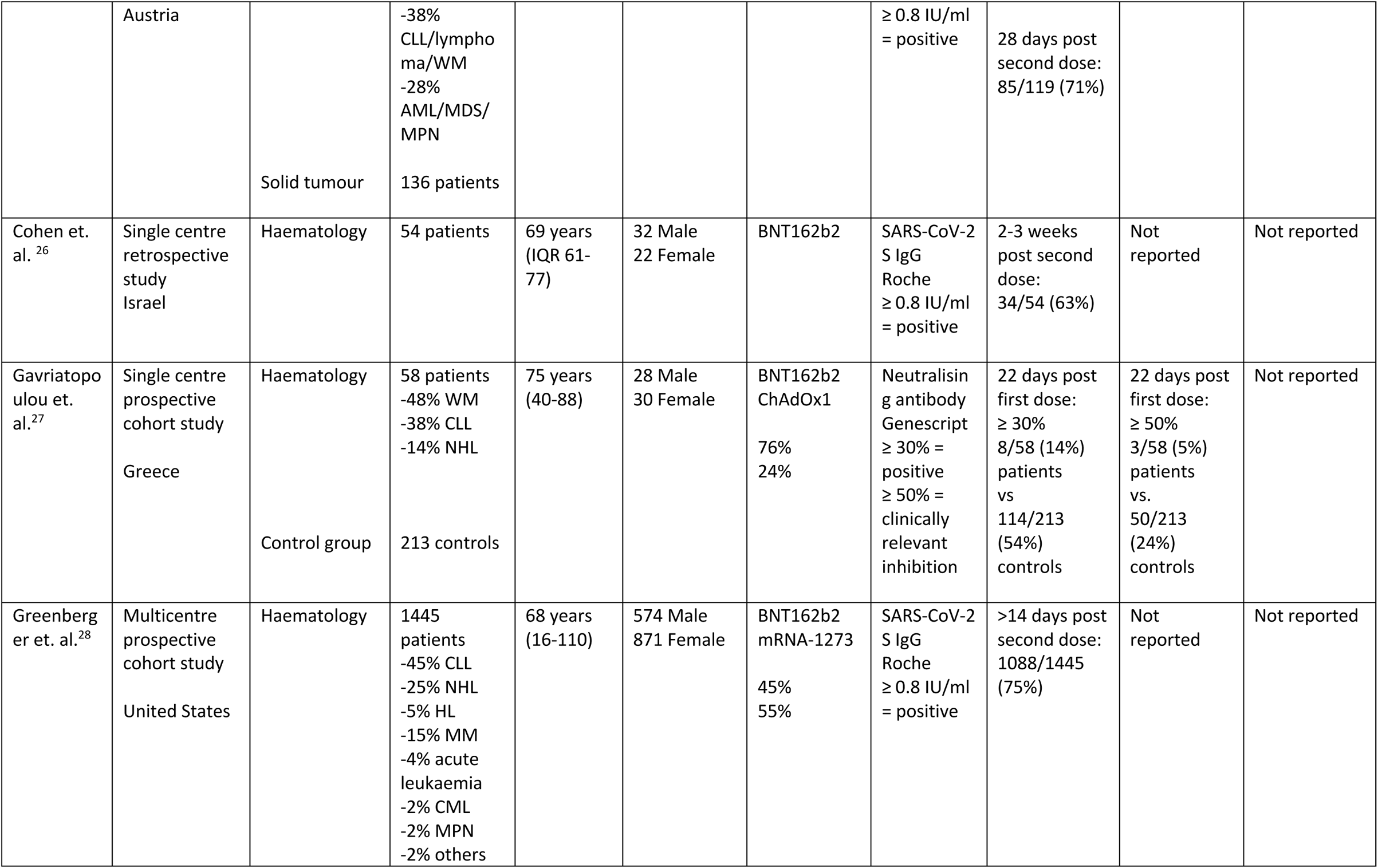
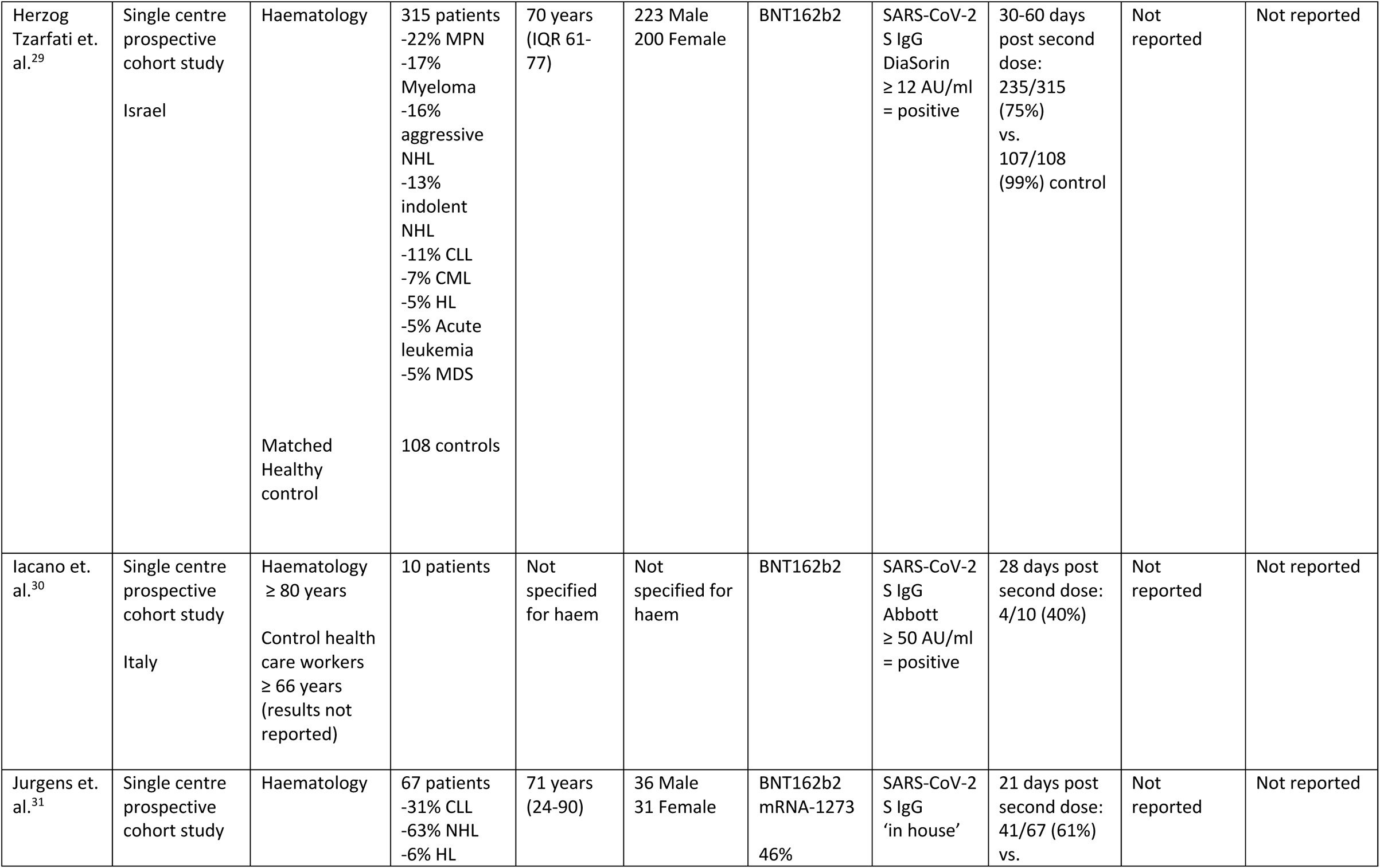
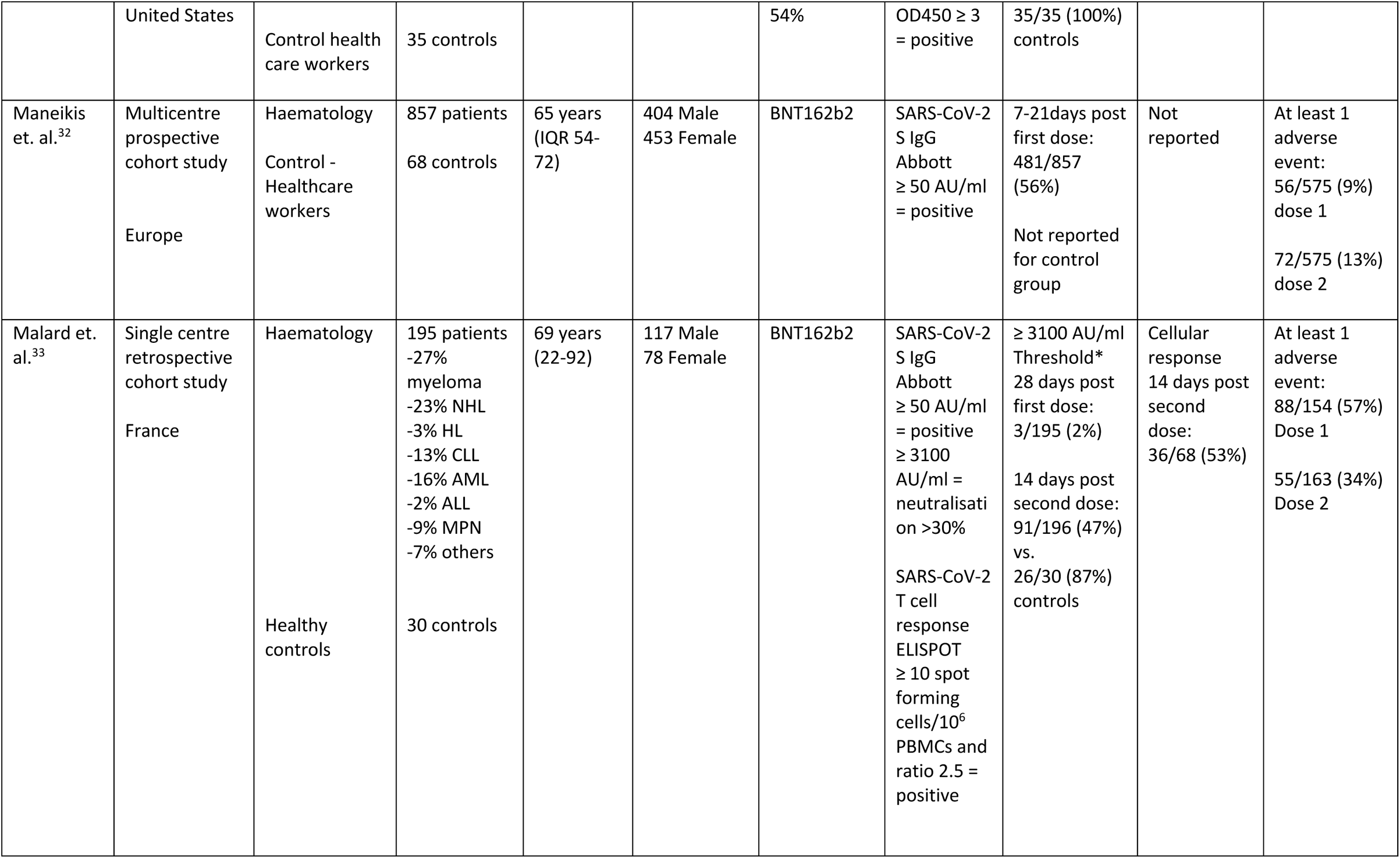
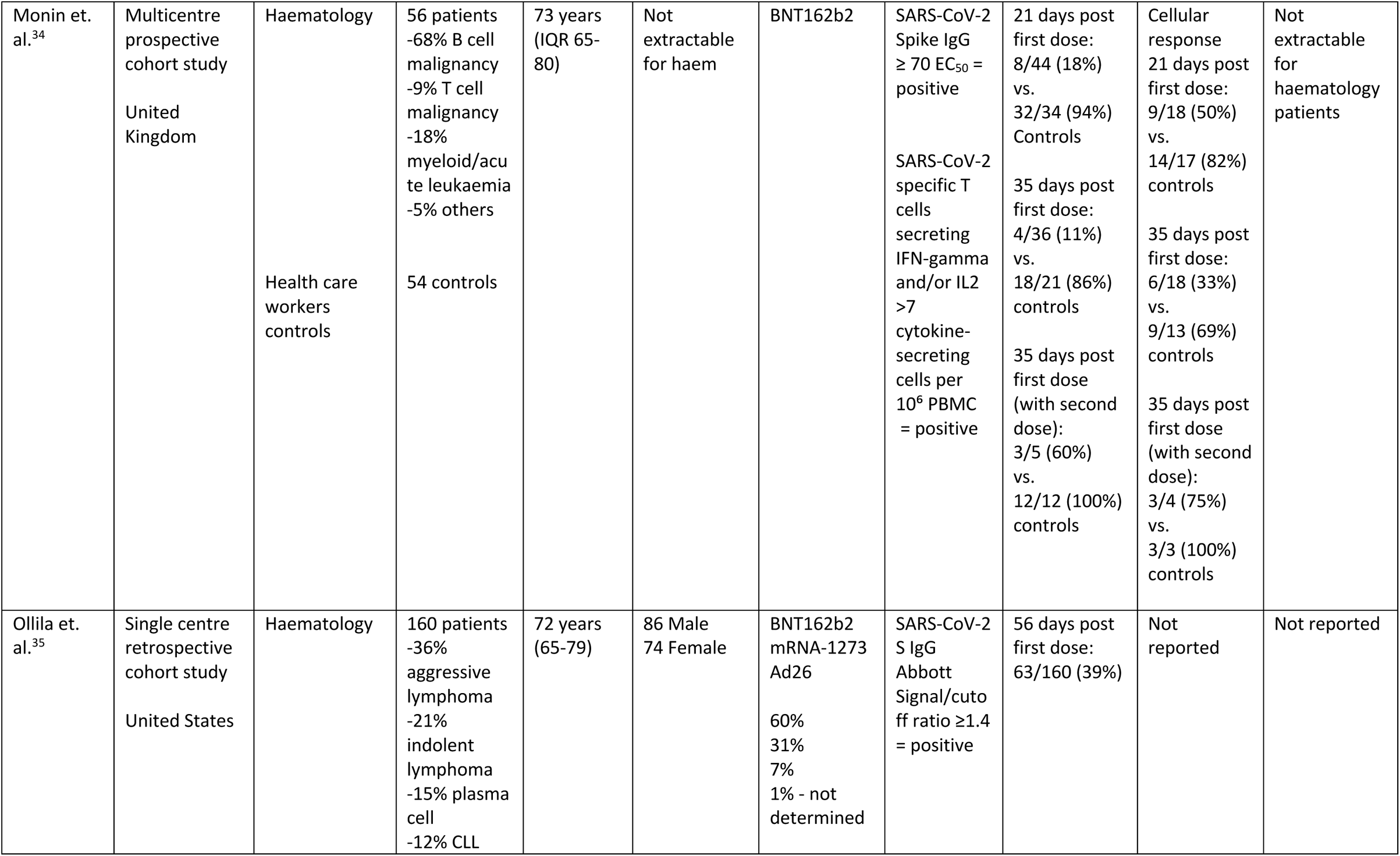
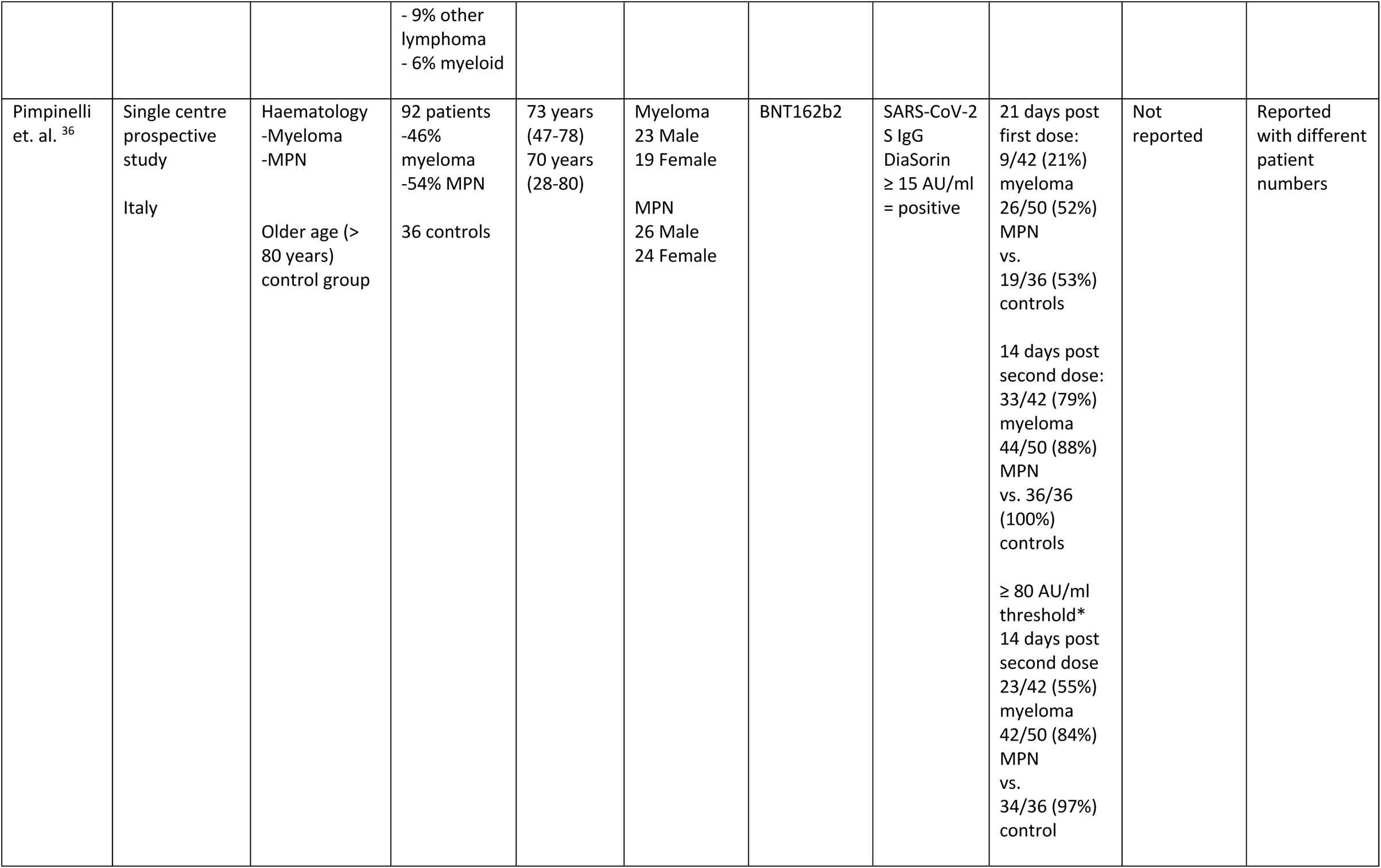
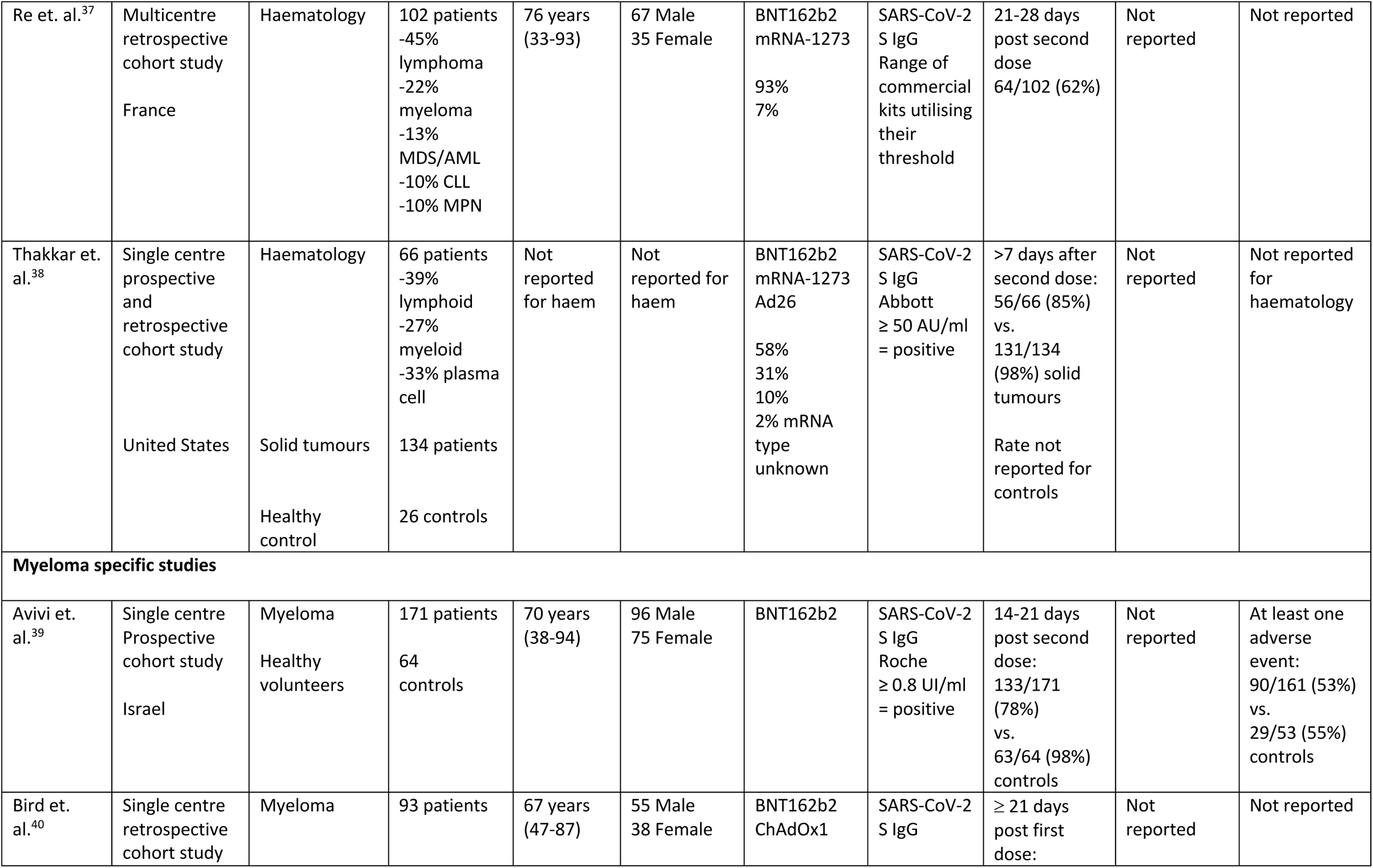
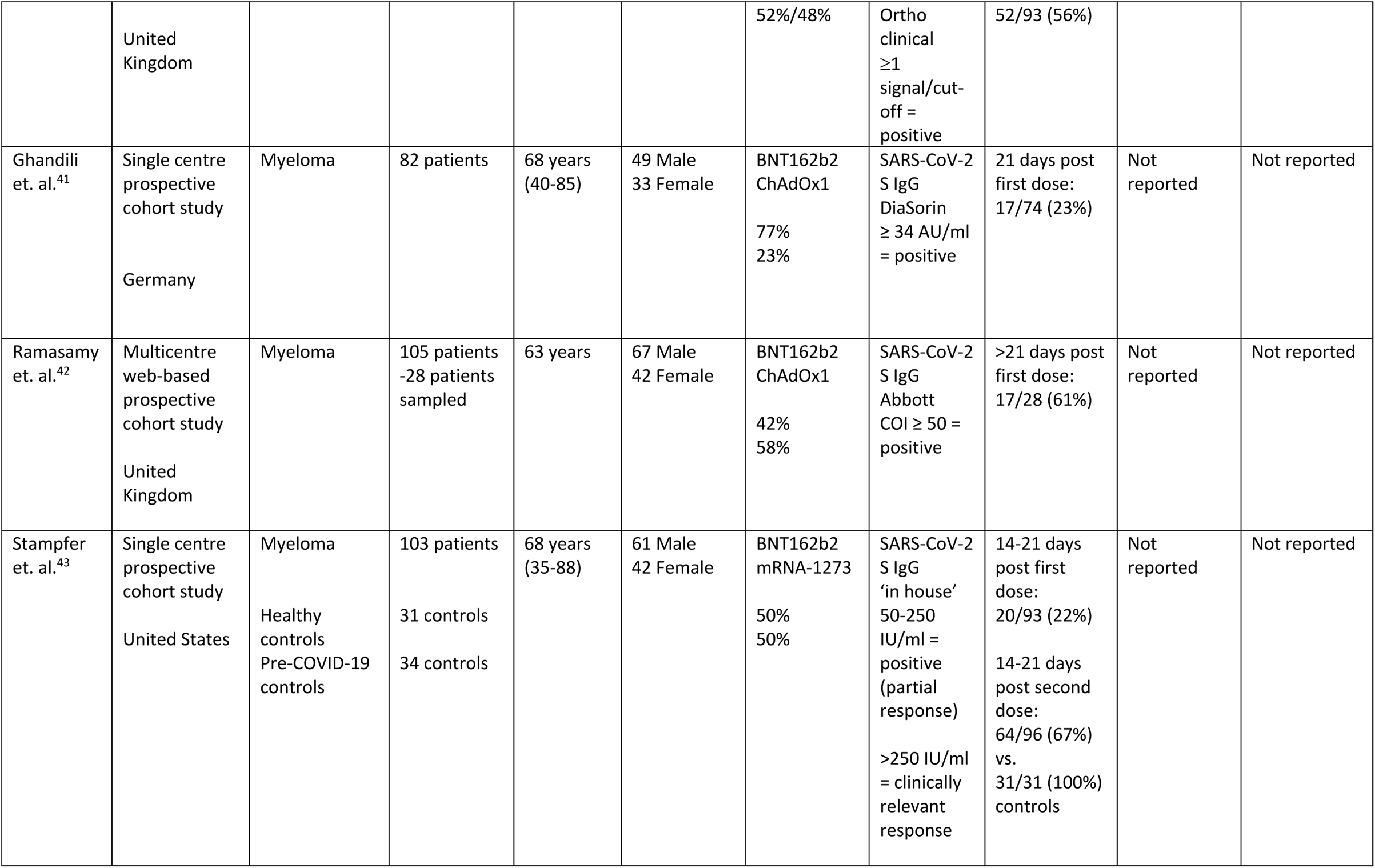
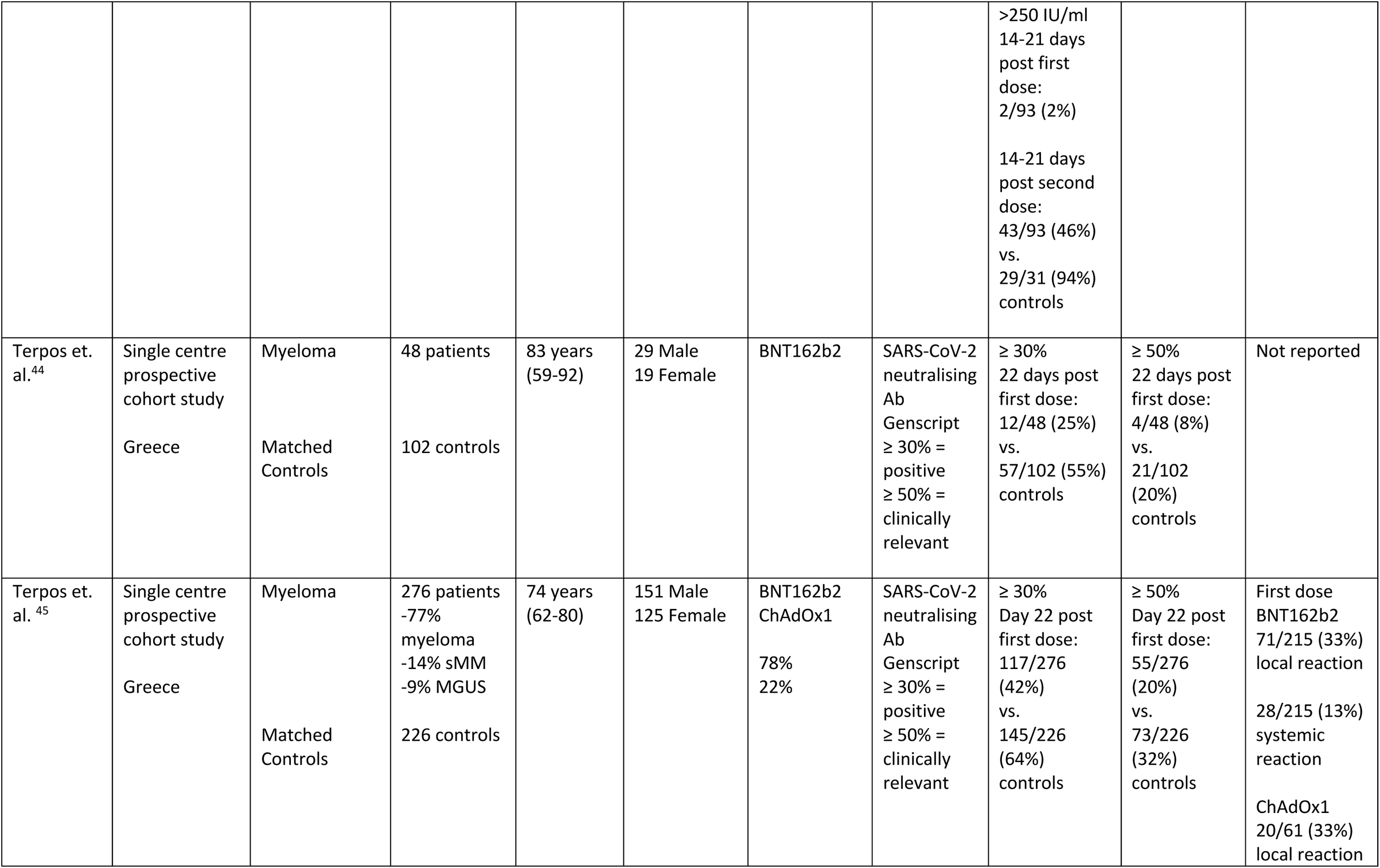
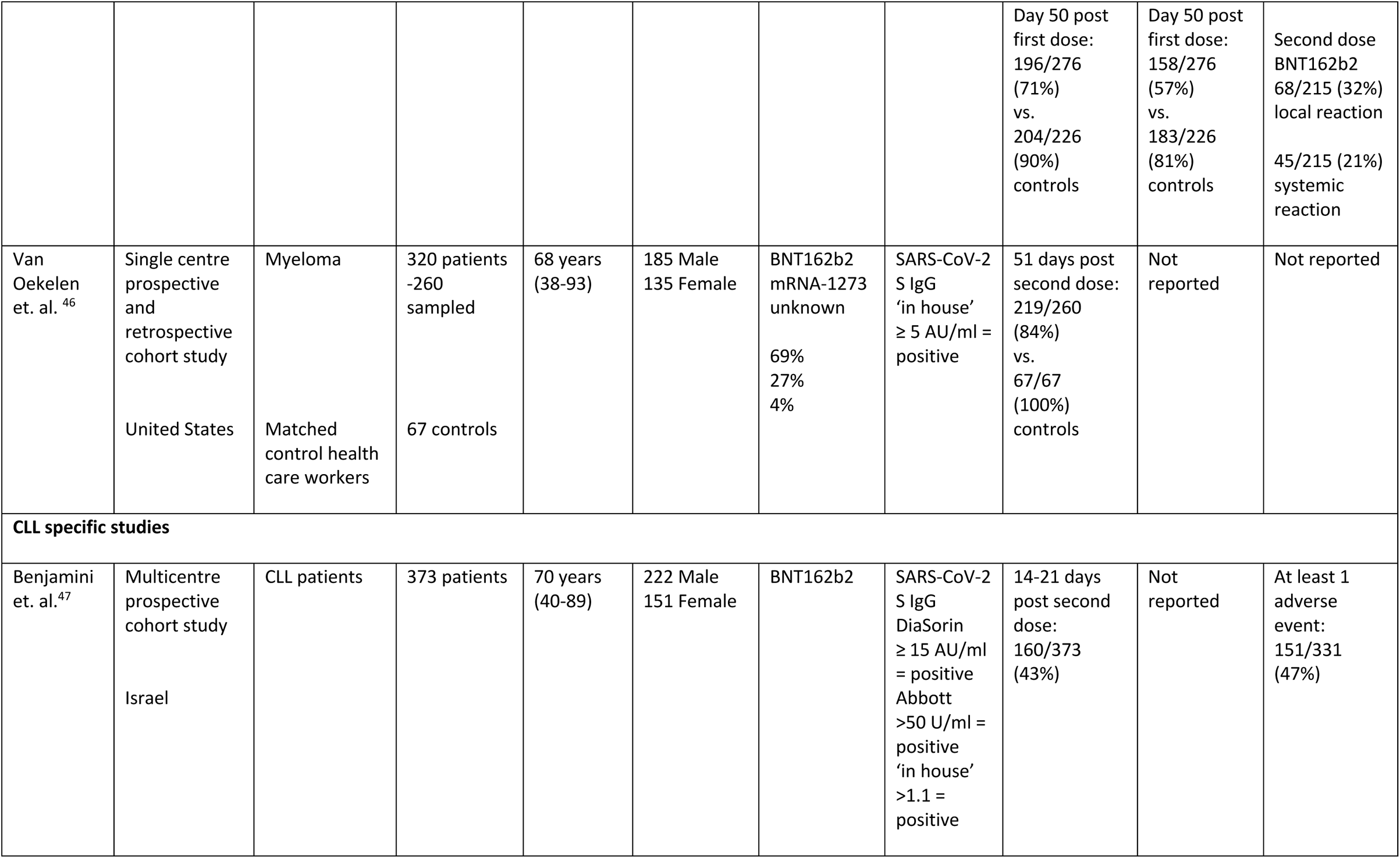
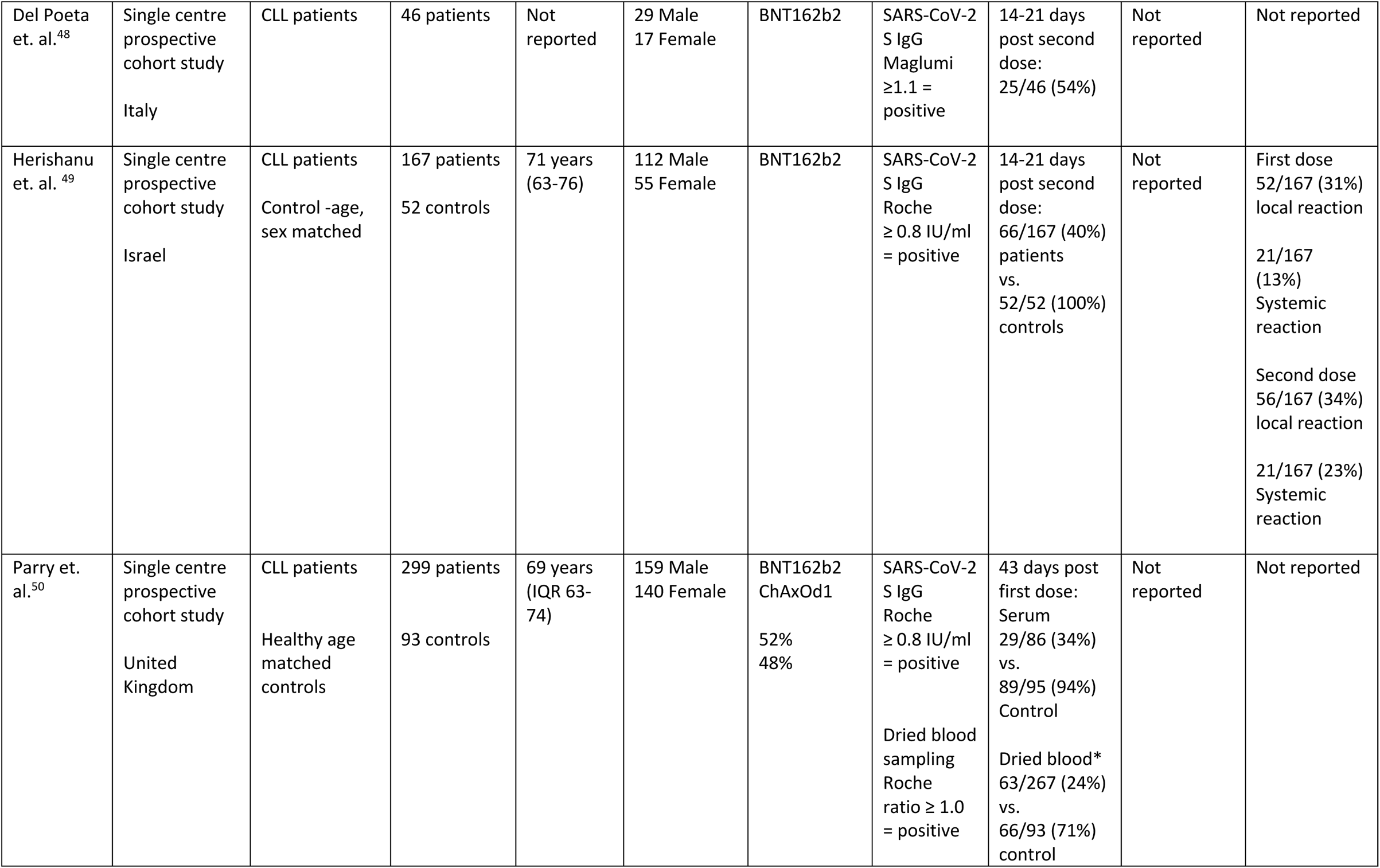
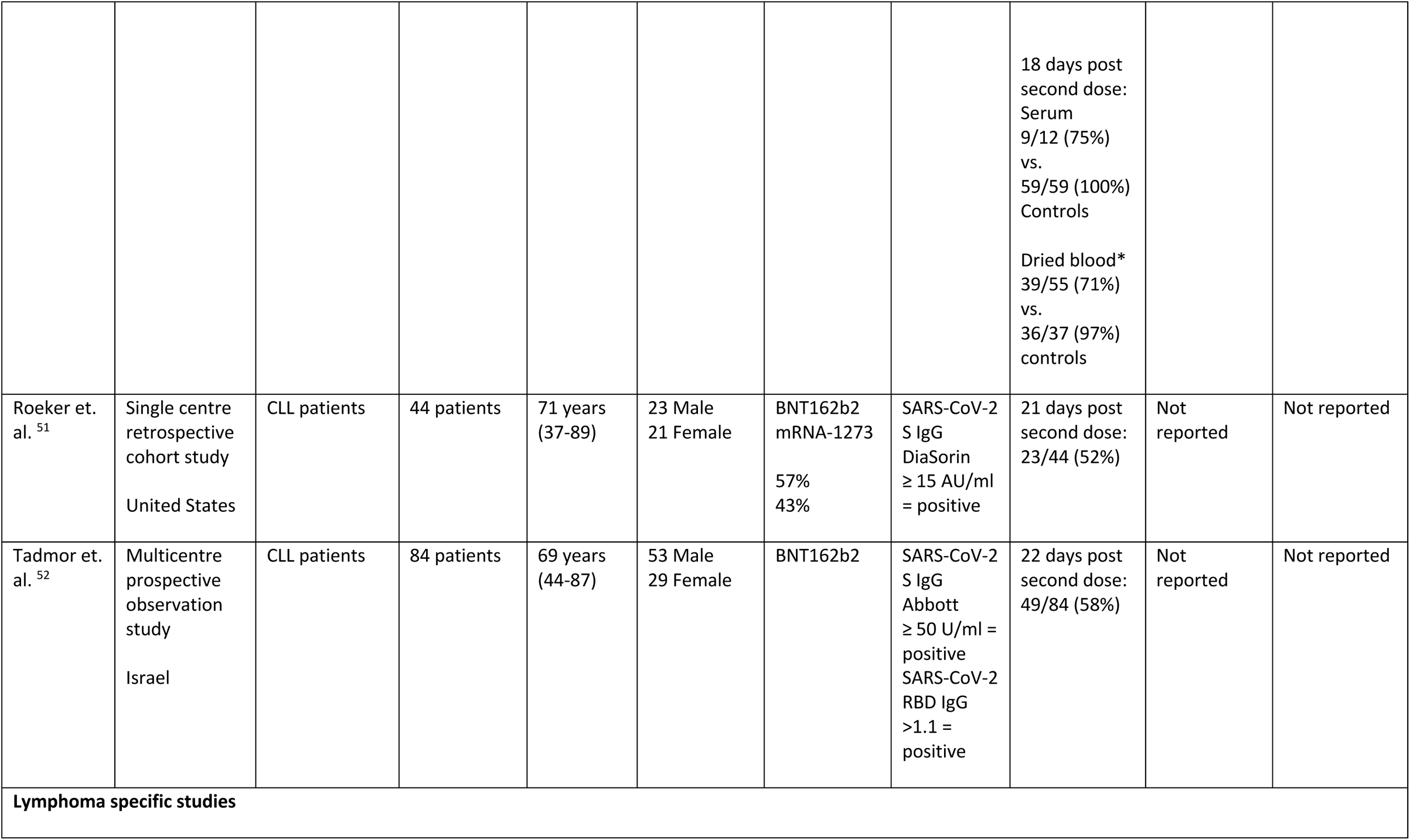
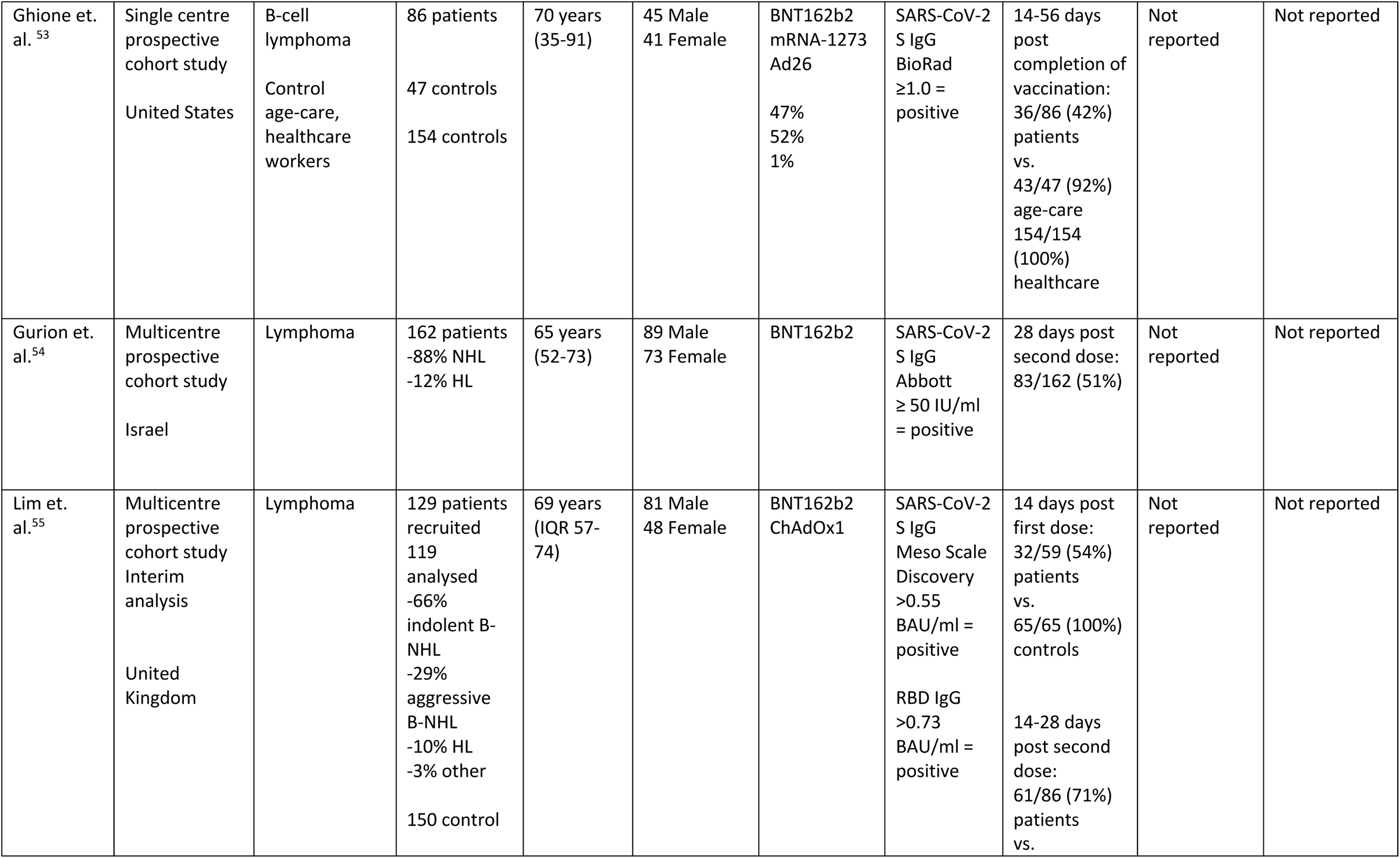
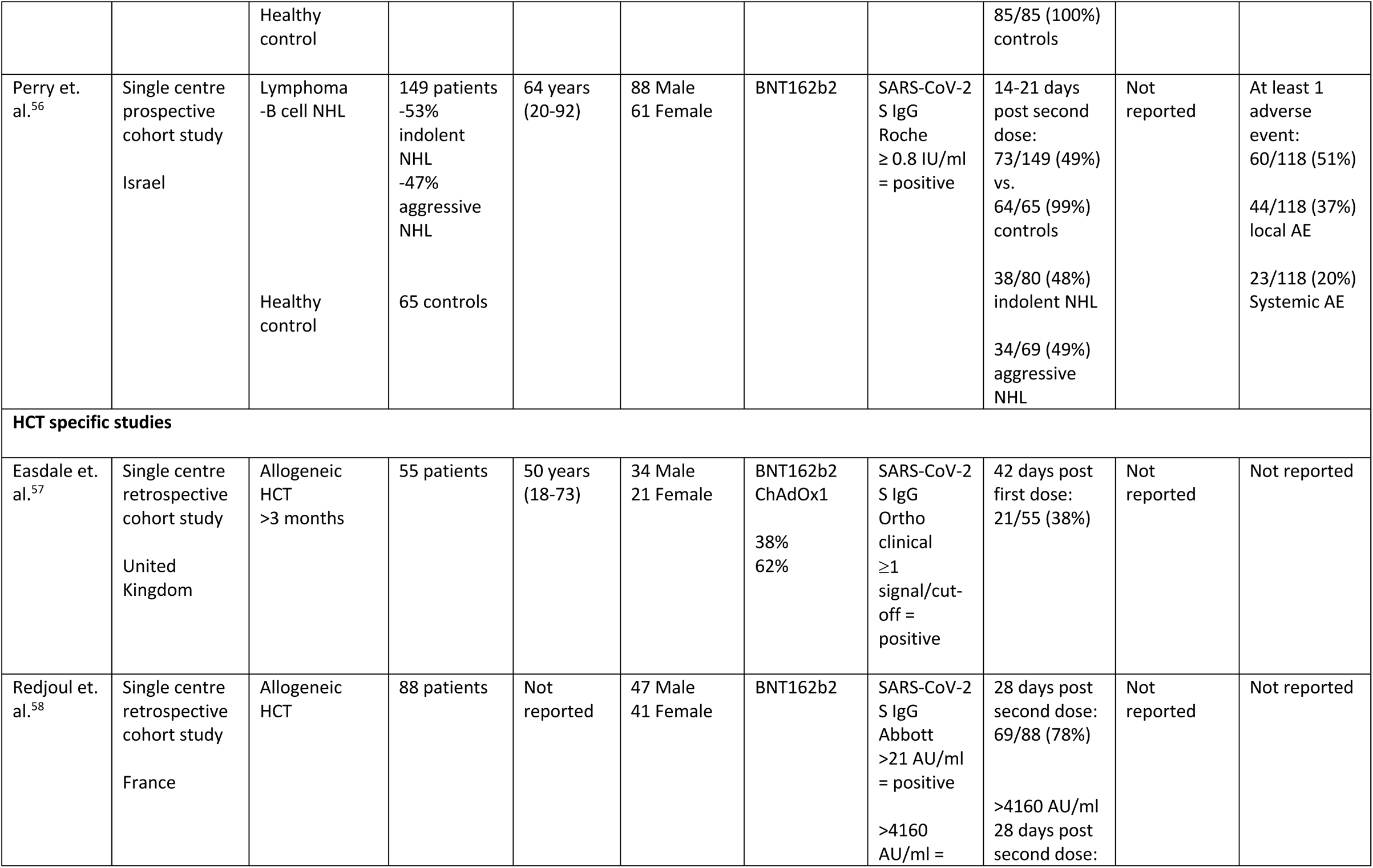
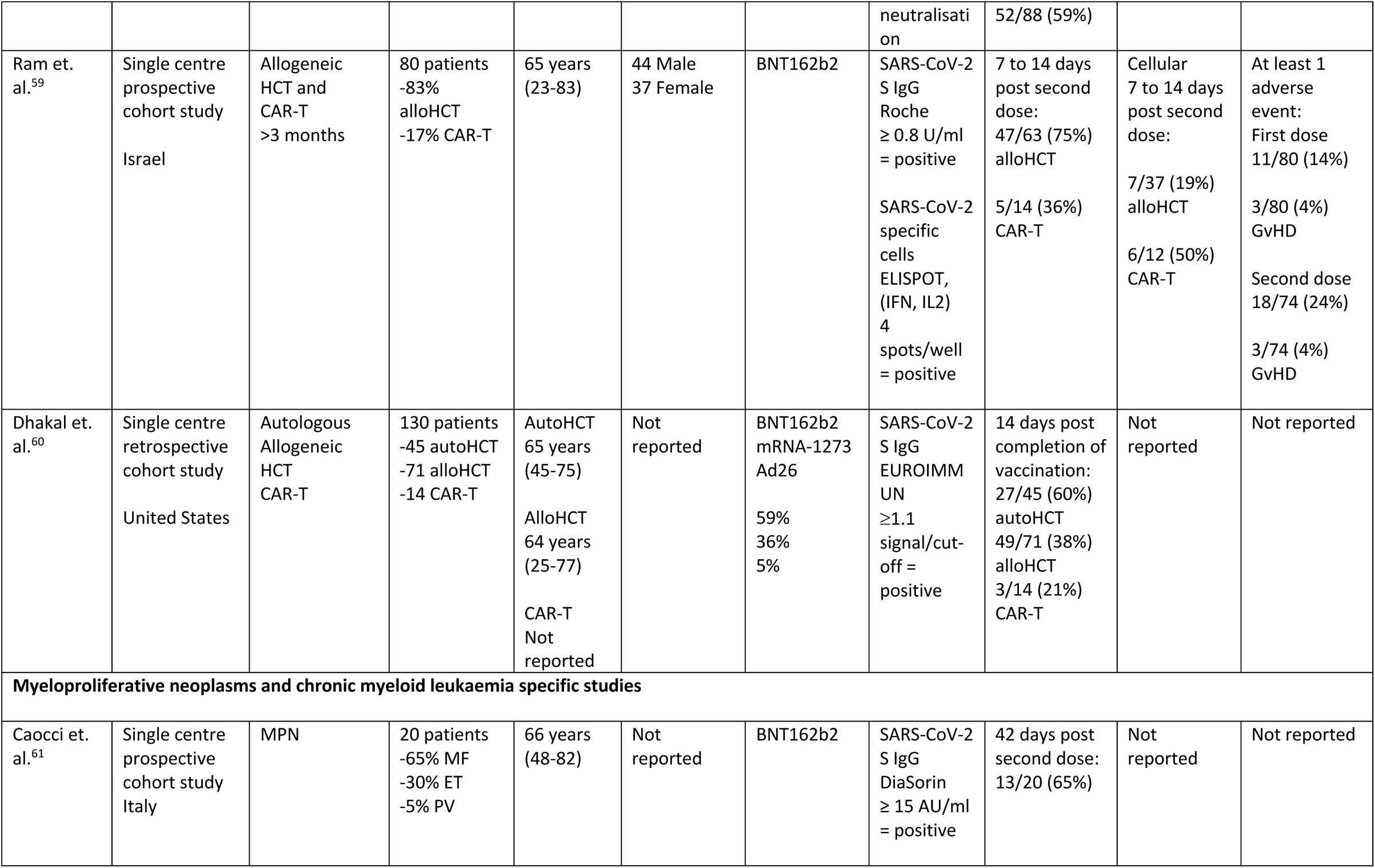
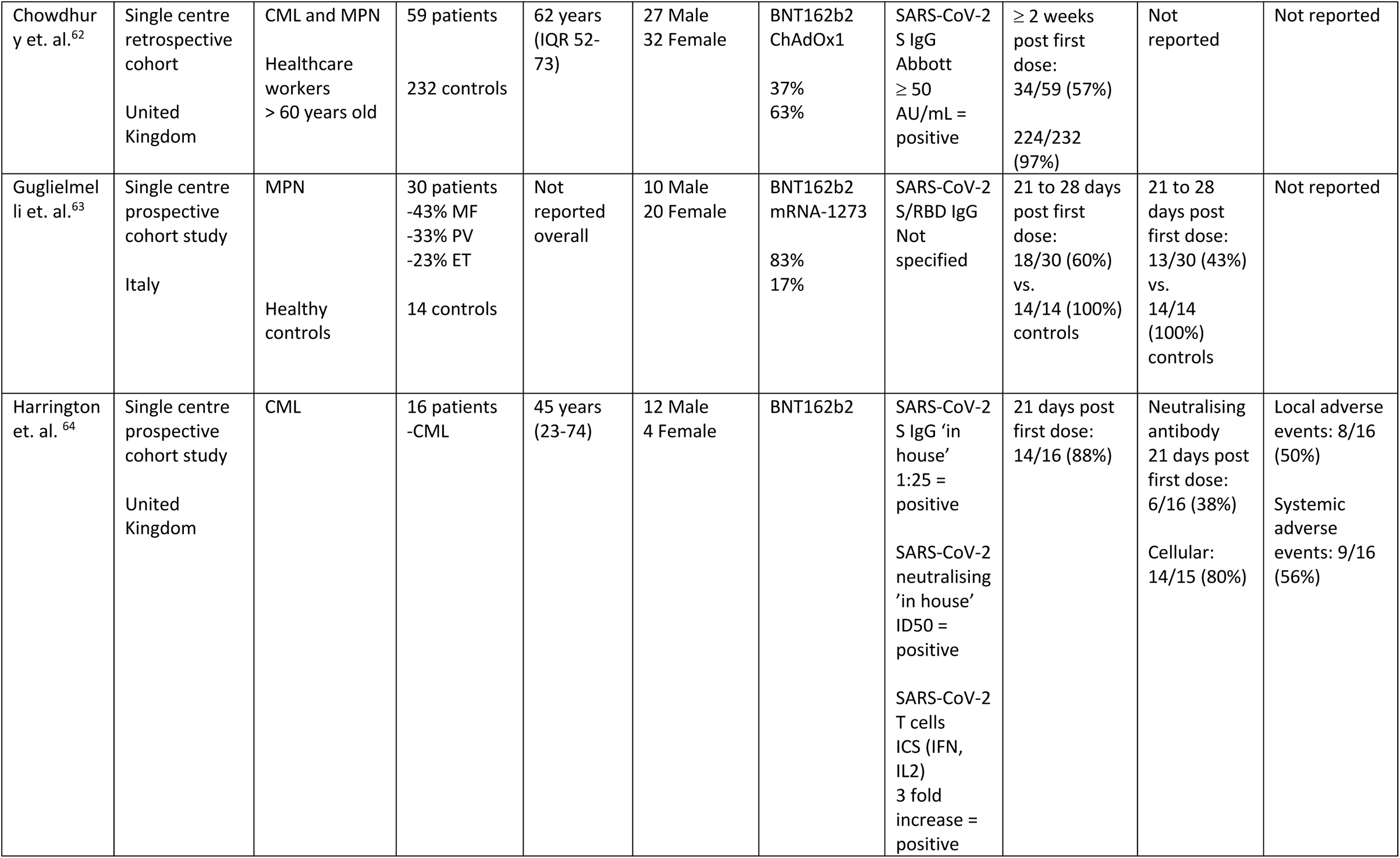
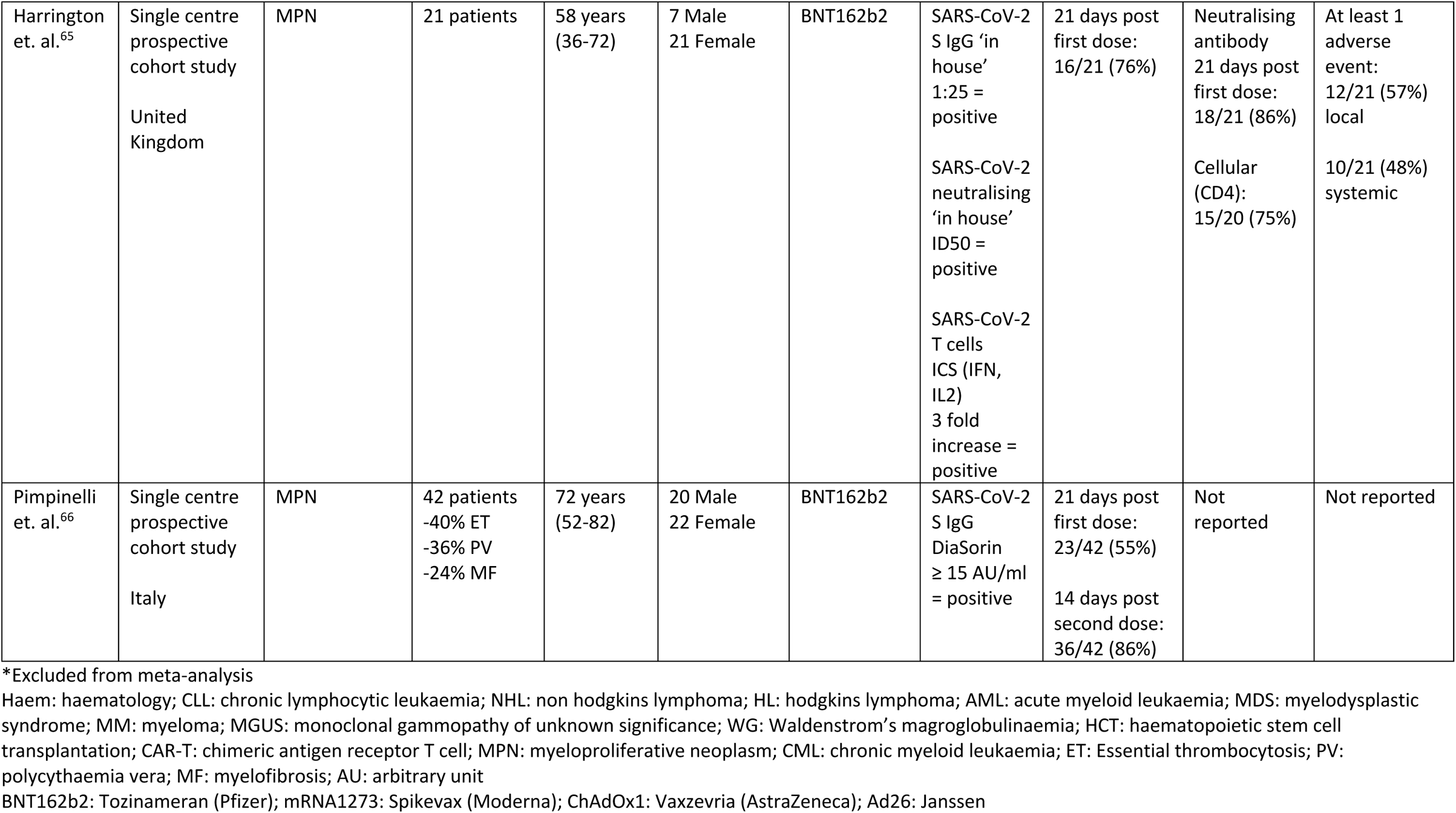
Summary of study characteristics and outcomes for patients with haematological malignancy by overall cohort and by specific underlying disease

### Risk of bias and quality assessment

Overall 23 studies (52%) were evaluated to be good or fair quality (Good = 11, Fair = 12) with low risk of bias whilst the remaining 21 studies were rated as poor quality (high risk of bias) predominantly due to lack of details regarding patient selection, demonstration that outcome of interest was not present at start of study, comparability of cohorts (design or analysis) and follow up duration. Risk of bias and quality assessment of studies are summarised in Supplementary Table 1.

### Seropositivity rates

There were 44 studies measuring humoral immune responses in patients with haematological malignancy. The majority of studies evaluated the use of BNT162b2 (Tozinameran, Pfizer) and utilised SARS-CoV-2 spike specific IgG levels above the threshold of detection (seropositivity). Several studies attempted to establish potential thresholds for clinical protection with higher antibody levels than the seropositivity threshold utilised. Malard et. al. correlated SARS-CoV-2 spike IgG levels with nAb levels and ≥3100 AU/mL on the Abbott assay was predictive of a nAb ≥ 30%^33^. Utilising this threshold, the rate of humoral response was 47% in haematology patients compared to 97% of healthy controls following second dose of the BNT162b2 vaccine^33^. Pimpinelli et. al. also evaluated humoral responses utilising a higher threshold of ≥ 80 AU/ml (Diasorin, Italy) and noted response rates of 55% for myeloma patients and 84% for MPN patients compared to 97% for healthy controls^36^. Other studies correlated SARS-CoV-2 IgG levels with plaque reduction neutralisation tests or with levels seen in clinical vaccination trials. Stampfer et. al. defined >250 IU/ml as a clinically relevant response which was attained by 45% of myeloma patients after two doses of a COVID-19 vaccine^43^. For HCT patients, Redjoul et. al. utilised a threshold of 4160 AU/ml and 59% of patients achieved this response after two doses^58^.

Overall pooled seropositivity rates in haematological malignancy were 61% and 67% in single arm and comparator studies respectively, following two doses of COVID-19 vaccination. Following a single dose, the pooled seropositivity rates were 51% and 37% in single arm and comparator studies respectively. Compared to healthy or aged matched controls, patients with haematological malignancy were less likely to achieve seropositivity with odds ratio (OR) of 0.05 (95% confidence interval [95% CI] 0.02-0.15, *p*<0.01) following two doses and OR of 0.10 (95% CI 0.04-0.29, *p*<0.01) following a single dose of COVID-19 vaccine. Heterogeneity was 86 and 88% respectively (*p*<0.01). Pooled seropositivity rates by disease type and vaccine doses are summarised in Table 2. Overall results were graded moderate quality due to statistical and clinical heterogeneity and the proportion of high risk of bias studies (48%).

**Table 2:** Summary of pooled seropositivity rates for patients with haematological malignancy and by underlying disease type and number of vaccine doses.

|  | Single arm studies<br>Pooled response rate<br>(95% CI) | Malignancy arm of<br>comparator studies<br>Pooled response rate<br>(95% CI) | Control arm of comparator<br>studies<br>Pooled response rate<br>(95% CI) | Intervention vs. control<br>cohort<br>Odds ratio (95% CI)<br>Heterogeneity<br>*p*-value |
| --- | --- | --- | --- | --- |
| **Overall: Patients with haematological malignancy** |  |  |  |  |
| Following second dose<br>4733 patients | 0.61 (0.54-0.69)<br>$I^2 = 92\%, p < 0.01$ | 0.67 (0.58-0.76)<br>$I^2 = 93\%, p < 0.01$ | 0.96 (0.91-1.00)<br>$I^2 = 84\%, p < 0.01$ | OR 0.05 (0.02-0.15)<br>$p < 0.01$<br>$I^2 = 88\%, p < 0.01$ |
| Following first dose<br>2331 patients | 0.51 (0.38-0.64)<br>$I^2 = 92\%, p < 0.01$ | 0.37 (0.23-0.51)<br>$I^2 = 90\%, p < 0.01$ | 0.78 (0.62-0.95)<br>$I^2 = 98\%, p < 0.01$ | OR 0.10 (0.04-0.29)<br>$p < 0.01$<br>$I^2 = 86\%, p < 0.01$ |
| **Myeloma** |  |  |  |  |
| Following second dose<br>1218 patients | 0.80 (0.64-0.95)<br>$I^2 = 85\%, p < 0.01$ | 0.76 (0.70-0.82)<br>$I^2 = 90\%, p < 0.01$ | 0.98 (0.95-1.00)<br>$I^2 = 71\%, p < 0.01$ | OR 0.09 (0.03-0.29)<br>$P < 0.01$<br>$I^2 = 49\%, p = 0.08$ |
| Following first dose<br>685 patients | 0.43 (0.18-0.68)<br>$I^2 = 91\%, p < 0.01$ | 0.29 (0.09-0.48)<br>$I^2 = 81\%, p < 0.01$ | 0.64 (0.42-0.87)<br>$I^2 = 76\%, p < 0.01$ | OR 0.23 (0.05-0.99)<br>$p = 0.05$<br>$I^2 = 49\%, p = 0.11$ |
| **Chronic lymphocytic leukaemia** |  |  |  |  |
| Following second dose | 0.51 (0.37-0.65) | 0.51 (0.34-0.68) | 1.00 (0.99-1.00) | OR 0.01 (0.01-0.03)<br>$p < 0.01$ |
| 1446 patients | $I^2 = 91\%, p < 0.01$ | $I^2 = 56\%, p = 0.06$ | $I^2 = 0\%, p = 0.07$ | $I^2 = 0\%, p = 0.9$ |
| Following first dose | Single study | 0.18 (0.00-1.00) | 0.92 (0.52-1.00) | OR 0.03 (0.00-0.80) |
| 111 patients | | $I^2 = 89\%, p < 0.01$ | $I^2 = 0\%, p = 0.32$ | $p = 0.05$<br>$I^2 = 0\%, p = 0.60$ |
| **Lymphoma** |  |  |  |  |
| Following second dose | 0.52 (0.28-0.75) | 0.55 (0.35-0.76) | 0.99 (0.98-1.00) | OR 0.02 (0.01-0.02) |
| 1227 patients | $I^2 = 97\%, p < 0.01$ | $I^2 = 84\%, p < 0.01$ | $I^2 = 0\%, p = 0.72$ | $p < 0.01$<br>$I^2 = 0\%, p = 1.0$ |
| Following first dose | No studies available | 0.33 (0.00-1.00) | 0.95 (0.09-1.00) | OR 0.01 (0.0-1.24) |
| 69 patients | | $I^2 = 93\%, p < 0.01$ | $I^2 = 71\%, p = 0.06$ | $p = 0.05$<br>$I^2 = 0\%, p = 0.7$ |
| **Haematopoietic stem cell transplant and cellular therapies** |  |  |  |  |
| Following second dose | 0.61 (0.42-0.80) | Single study | Single study | Single study |
| 401 patients | $I^2 = 88\%, p = 0.05$ | | | |
| Following first dose | Single study | No studies available | No studies available | No studies available |
| 21 patients |  |  |  |  |
| **Acute leukaemia and Myelodysplastic syndrome** |  |  |  |  |
| Following second dose | 0.93 (0.80-1.00) | Single study | Single study | Single study |
| 113 patients | $I^2 = 29\%, p = 0.24$ | | | |
| Following first dose<br>13 patients | Single study | Single study | Single study | Single study |
| **Myeloproliferative neoplasm and chronic myeloid leukaemia** |  |  |  |  |
| Following second dose<br>281 patients | 0.88 (0.72-1.00)<br>$I^2 = 70\%, p=0.01$ | 0.87 (0.81-0.92)<br>$I^2 = 0\%, p=0.83$ | 0.99 (0.98-1.00)<br>$I^2 = 0\%, p=0.90$ | OR 0.07 (0.0-1.55)<br>$p = 0.06$<br>$I^2 = 0\%, p=0.77$ |
| Following first dose<br>222 patients | 0.71 (0.30-1.00)<br>$I^2 = 76\%, p=0.01$ | 0.54 (0.37-0.71)<br>$I^2 = 0\%, p=0.46$ | 0.85 (0.51-1.00)<br>$I^2 = 90\%, p<0.01$ | OR 0.13 (0.01-1.71)<br>$p=0.09$<br>$I^2 = 88\%, p<0.01$ |

### Patients with myeloma

There were 8 dedicated studies and 9 other studies that reported immune response rates in myeloma patients (Supplementary Table 2). For the two studies by Terpos et. al, only nAb at two levels were reported and thresholds of ≥30% and ≥50% were utilised to define seropositivity and rate of positive nAb respectively^44,45^. There were 1218 myeloma patients who received 2 doses of vaccine and seropositivity rates were 76-80%. Seropositivity rates were 29-43% following 1 dose of COVID-19 vaccine in 685 patients. The OR for achieving seropositivity was 0.09 (95% CI 0.03-0.29, *p*<0.01) in myeloma patients compared to healthy control group following 2 doses and 0.23 (95% CI 0.05-0.99*, p*=0.05) following 1 vaccine dose (Table 2).

### Patients with CLL

A total of 1557 CLL patients in 12 studies (6 CLL specific, 6 haematology) were included in this review (Supplementary Table 3). Pooled seropositivity rates were 51% in 1446 CLL patients following 2 doses of COVID-19 vaccine. Rate of seropositivity was 18% following 1 dose of COVID-19 vaccine and it was 37% in one single arm study by Ollila et. al^35^. ORs for seropositivity were 0.01 (95% CI 0.01-0.03, *p*<0.01) and 0.03 (95% CI 0.00-0.80, *p*=0.05) for CLL patients following 2 doses and 1 dose of vaccine compared to healthy controls.

### Patients with lymphoma

For 1296 patients with lymphoma across 11 studies included (Supplementary Table 4), pooled seropositivity rates were 52-55% following 2 doses and 33% following 1 dose of COVID-19 vaccine. Compared to healthy cohort, ORs for achieving seropositivity were 0.01 to 0.02 for lymphoma patients (Table 2).

### Patients following HCT and CAR-T

A total of 6 studies included in this review reported on immune responses following HCT and CAR-T therapy (Supplementary Table 5). Two studies involved allogeneic HCT (alloHCT) patients^57,58^, one included alloHCT and CAR-T treated patients^59^ and the other study included all HCT and CAR-T patients^60^. Two studies were larger studies of haematology patients with subset of HCT and CAR-T patients^28,29^. 422 patients were included consisting of 401 patients who had 2 doses and the remainder received 1 dose of COVID-19 vaccine. Pooled seropositivity rate was 61% following 2 COVID-19 vaccine doses. For patients following alloHCT the rate was 74% while following CAR-T, it was 31%. In a single study with a healthy control cohort, seropositivity rate was 81% in autologous HCT patients compared to 99%^29^. In the only study of immune response following one COVID-19 vaccine dose, rate of seropositivity was 38% in alloHCT patients^57^.

### Patients with acute leukaemia and MDS

There were no dedicated studies of COVID-19 vaccination in patients with acute leukaemia. A subset of 126 patients with acute leukaemia and MDS were reported in 6 studies (Supplementary Table 6) and seropositivity rate was 93% following 2 doses of COVID-19 vaccine. In a single study, the seropositivity rate was 80% in acute leukaemia, 94% in MDS patients compared to 99% in the control group^29^. No patients with acute leukaemia mounted an immune response following a single vaccine dose compared to 86% of controls^34^.

### Patients with MPN and CML

Of 12 studies encompassing 503 MPN and CML patients (281 following 2 doses, 222 following a single dose), rates of seropositivity were 87-88% following 2 doses and 54-71% following 1 dose of vaccine. Compared to a healthy patient cohort, the OR was 0.07 (95% CI 0.0-1.55, *p*=0.06) of MPN/CML patient achieving seropositivity with 2 doses of COVID-19 vaccine and 0.13 (95% CI 0.01-1.71, *p*=0.09) after a single dose.

### Rates of neutralising antibody, cellular response and adverse events

Only 6 studies (14%) reported nAb responses and 4 studies (9%) reported cellular responses. In the only study on nAb following 2 COVID-19 vaccine doses, 71% of myeloma patients achieved a positive nAb response compared to 90% of the control group^45^. Following 1 dose of COVID-19 vaccine, the overall pooled rate of a positive nAb response was 18% for all patients with haematological malignancy (Table 3). The pooled rate of 2 single arm studies of MPN patients was 63%. For cellular responses following COVID-19 vaccination, the rate of achieving a positive response was 40-75% for all patients following 2 doses and 33-86% following a single dose. In a single study of alloHCT and CAR-T patients, cellular response rates were 19% and 50% respectively following 2 doses^59^. In MPN patients, this rate was 86% following one dose^64,65^. In 10 studies (22%) the rate of at least one systemic or local adverse event was reported. Overall, the pooled rate of at least one adverse event was 36% following 2 doses and 39% following a single dose (Table 3).

**Table 3:** Summary of pooled rates of positive neutralising antibody, cellular responses and adverse events

|  |  |  |  |
| --- | --- | --- | --- |
|  | Positive neutralising antibody response | Positive cellular response | At least 1 adverse event rate |
|  | Pooled response rate (95% CI) | Pooled response rate (95% CI) | Pooled response rate (95% CI) |
| **Overall: Patients with haematological malignancy** |  |  |  |
| Following second dose | Malignancy arm of comparator studies: 0.57 (Single study) | Single arm: 0.40 (0.00-1.00)<br>Malignancy arm of comparator studies: 0.75 (single study) | 0.36 (0.24-0.48)<br>$I^2 = 97\%, p < 0.01$ |
| Following first dose | Single arm: 0.63 (0.00-1.00)<br>Malignancy arm of comparator studies: 0.18 (0.00-0.44) | Single arm: 0.86 (0.00-1.00)<br>Malignancy arm of comparator studies: 0.33 (single study) | 0.39 (0.18-0.60)<br>$I^2 = 97\%, p < 0.01$ |

### Subgroup analysis

Study characteristics and outcomes of studies included in subgroup analyses are summarised in Supplementary Tables 8-12. Vaccination with 2 vaccine doses during active therapy was associated with seropositivity rates of 28% compared to 62% off therapy with OR 0.21 (95% CI 0.06-0.67, *p*=0.02). Lower seropositivity rates (19% vs. 61%) were reported with vaccination during or within 12 months of CD-20 antibody therapy compared to vaccination 12 or more months after completion of therapy. Use of targeted therapy was associated with a pooled seropositivity rate of 35% following 2 doses of COVID-19 vaccine. Seropositivity rates did not differ by timing of vaccination in relation to HCT (66% vs 68%) or vaccine type (62% vs 68%, 95% vs 91%) following 1 or 2 doses. Table 4 summarises the seropositivity rates by each subgroup analysed.

**Table 4:** Summary of subgroup analysis by active treatment, timing of CD20 antibody therapy, haematopoietic stem cell transplantation, receipt of targeted therapies and type of COVID-19 vaccine

|  | Intervention arm<br>Pooled response rate (95% CI)<br>Heterogeneity<br>*p*-value | Control arm<br>Pooled response rate (95% CI)<br>Heterogeneity<br>*p*-value | Intervention vs. control cohort<br>Odds ratio (95% CI)<br>Heterogeneity<br>*p*-value |
| --- | --- | --- | --- |
| **Active therapy vs. no active therapy** |  |  |  |
| Following second dose | 0.28 (0.08-0.48)<br>$I^2 = 83\%, p < 0.01$ | 0.62 (0.39-0.86)<br>$I^2 = 94\%, p < 0.01$ | OR 0.21 (0.06-0.67)<br>$p = 0.02$<br>$I^2 = 75\%, p < 0.01$ |
| Following first dose | 0.42 (0.09-0.75)<br>$I^2 = 49\%, p = 0.08$ | 0.81 (0.66-0.95)<br>$I^2 = 0\%, p = 0.62$ | OR 0.19 (0.03-1.19)<br>$p = 0.06$<br>$I^2 = 24\%, p = 0.27$ |
| **CD-20 antibody therapy within 12 months vs. CD-20 therapy 12 or more months** |  |  |  |
| Following second dose | 0.19 (0.00-0.50)<br>$I^2 = 88\%, p < 0.01$ | 0.61 (0.41-0.82)<br>$I^2 = 88\%, p < 0.01$ | OR 0.08 (0.01-0.59)<br>$p = 0.02$<br>$I^2 = 57\%, p = 0.04$ |
| Following first dose | Single study | Single study | Single study |
| **Targeted therapy** |  |  |  |
| Following second dose | 0.35 (0.19-0.52)<br>$I^2 = 94\%, p < 0.01$ | No control | No analysis |
| Following first dose | Single study | No control | No analysis |

| **Haematopoietic stem cell transplant or cellular therapy within 12 months vs. 12 or more** |  |  |  |
| --- | --- | --- | --- |
| Following second dose | 0.66 (0.43-0.88)<br>$I^2 = 0\%, p=0.63$ | 0.68 (0.40-0.95)<br>$I^2 = 45\%, p=0.16$ | OR 0.96 (0.11-8.74)<br>$p=0.94$<br>$I^2 = 36\%, p=0.21$ |
| Following first dose | Single study | Single study | Single study |
| **Vaccine type BNT162b2 vs. non-BNT162b2** |  |  |  |
| Following second dose | 0.95 (0.39-1.00)<br>$I^2 = 66\%, p=0.09$ | 0.91 (0.62-1.00)<br>$I^2 = 0\%, p=0.44$ | OR 1.93 (0.00- >1)<br>$p=0.69$<br>$I^2 = 71\%, p=0.06$ |
| Following first dose | 0.62 (0.29-0.95)<br>$I^2 = 52\%, p=0.12$ | 0.68 (0.24-1.00)<br>$I^2 = 76\%, p=0.02$ | OR 0.80 (0.42-1.51)<br>$p=0.26$<br>$I^2 = 0\%, p=0.82$ |

### Sensitivity analysis

Exclusion of studies that were assessed as poor quality and high risk of bias did not alter the pooled seropositivity rates (single arm) but in comparator studies, OR was now 0.31 (95%CI 0.13-0.78, *p*=0.03) following 1 dose instead of 0.10 (Supplementary Table 13).

## Discussion

In this systematic review and meta-analysis of over 7000 patients with haematological malignancy, rate of seropositivity was 61-67% following 2 doses and 37-51% following 1 dose of COVID-19 vaccination. Compared to age matched and non-matched healthy controls primarily healthcare workers, odds of achieving seropositivity were significantly lower by 95% and 90% following 2 doses and 1 dose of COVID-19 vaccination respectively. Statistical heterogeneity was substantial at over 80% and studies were also clinically heterogenous due to the variety of underlying haematological malignancies, lack of standardised laboratory platforms by which to measure immune response to vaccination and variable follow up periods. Reassuringly, reported rates of at least one adverse effect following vaccination were lower at 36% compared to rates reported in clinical trials of the general population^10,13^.

Amongst different haematological malignancies, pooled seropositivity rates following 2 doses were highest at close to 90% in patients with acute leukaemia, MDS and MPN and the lowest at 51% in CLL patients. Patients with CLL respond poorly to vaccination, especially to new or novel antigens compared to recall antigens from previous infections or vaccination^67^. Poor response is compounded by use of B-cell depleting and targeted therapies such as CD-20 monoclonal antibodies and BTKi^67^. Immune response to other vaccines including seasonal influenza vaccine is negatively impacted by these therapies and the poor response persists for at least 6 to 12 months following cessation of therapy^68^.

A similar negative impact of treatment on COVID-19 vaccine responses was noted in subgroup analysis of published studies. Rates of seropositivity were lower in the setting of active treatment (28%) and the lowest response rates of 19%, 35% and 31% were reported in the setting of current or CD-20 monoclonal antibody therapy within 12 months, targeted therapies and following CAR-T cell therapy. Seropositivity rates were two to three times higher at 61 and 62%% when patients were vaccinated when not on active therapy or 12 or more months following completion of CD-20 monoclonal antibody respectively.

Unvaccinated CLL patients have a high burden of morbidity from COVID-19 infection with close to 90% of patients requiring hospital admission, 35% intensive care unit admission and mortality rates^4^. Yet, CLL patients have poor humoral responses to vaccination at 18% following 1 dose and 51% following 2 doses. In CLL patients, immune suppression from underlying disease and ongoing treatments such as anti-CD20 monoclonal antibodies and BTKi continue to pose a risk for SARS-CoV-2 infection and concurrently limit protective responses from vaccination. Although vaccination remains highly recommended, new strategies are required to further improve immune responses in this high-risk patient population. In other immune compromised groups, the use of an additional dose of COVID-19 vaccine improved serological response rates by 37% and use of heterologous vaccination schedules (mixing vaccine types) appear promising^69–71^. International health bodies now recommend third COVID-19 vaccine doses in immune compromised patients including haematological malignancy^72,73^. Heterologous prime-boost or use of high dose vaccine formulations have been utilised in randomised trials of seasonal influenza vaccination in haematology and HCT patient with mixed success^17,74^. In addition, other preventative approaches such as the use of anti-SARS-CoV-2 monoclonal antibody therapy as pre exposure (NCT04625725) and post exposure prophylaxis require further evaluation in vulnerable patient groups who respond poorly to vaccination^75^.

Serological responses are classically utilised as surrogate end points for clinical efficacy in clinical trials of vaccination in patients with haematological malignancy^15–17^. Serological thresholds for protection (seroprotection) following vaccination have been established for infections such as seasonal influenza^17^. In the majority of studies included in this review, however, the outcome of interest was seropositivity as defined by antibody levels above the detection threshold. This is not equivalent to seroprotection as thresholds have not been standardised nor established across the variety of commercial and research platforms utilised by these studies. Some authors such as Redjoul, Stampfer and Malard et. al. have attempted to define serological thresholds that correlate with nAb and clinical protection^33,43,58^. Unsurprisingly a lower proportion of patients (by 20-35% compared to pooled rates) achieve these higher antibody thresholds^33,43,58^. For serological response measurement to guide clinical management of COVID-19 vaccination, further work is required to achieve harmonisation across testing platforms and to derive and validate thresholds that correlate with clinical protection.

Measurement of serological responses only offers a glimpse of the potential breadth of immune responses to COVID-19 vaccination. Both nAb and cellular responses to vaccination play complementary and vital roles in protection against COVID-19 and remain under reported^76–78^. As reported in 14% of studies, at least 18% of haematology patients achieved a positive nAb response. Positive cellular response rates were at least 15% higher than nAb responses following 1 or 2 vaccine doses. While serological responses have been utilised as surrogate endpoints in vaccination studies of immune compromised patients, more larger studies are required to identify new immune markers for vaccine response and to determine efficacy of vaccination.

There are several limitations to this review. In particular the moderate quality of findings due to significant clinical and statistical heterogeneity of included studies and the proportion of poorer quality studies. In line with other established studies of vaccination in haematology patients, only immune response data was analysed as clinical efficacy data was limited.

In conclusion, this systematic review and meta-analysis has comprehensively summarised the latest data on response to COVID-19 vaccination in patients with haematological malignancy. Overall, seropositive rates were reasonable at 67% following two doses of vaccination respectively. Higher risk patient groups were identified, namely patients with CLL and patients receiving active therapy including targeted and CD-20 monoclonal therapies. New approaches to high risk patients who are poor responders to vaccination are urgently required.

## Supporting information

PRISMA checklist

## Data Availability

All data produced in the present study are available upon reasonable request to the authors

## Acknowledgement

B.W.T is supported by the Australian Government Medical Research Future Fund Investigator Fellowship. M.A.S is supported by the National Health and Medical Research Council Investigator fellowship.

## Authorship contributions

B.W.T coordinated the study, performed abstract and full text screening, data extraction and manuscript writing. J.T performed abstract and full text screening and data extraction. J.C. developed the study protocol and performed data extraction. S.L. developed the search strategy. T.S. conducted the statistical analysis. Z.N and M.A.S conducted the risk of bias assessment. All authors reviewed and revised the manuscript.

## Conflict of interest

M.A.S. has received research funding and honoraria from Pfizer, Merck Sharp and Dohme and research funding from Gilead. B.W.T has received research funding and honoraria from CSL-Behring, Merck Sharp and Dohme and Sanofi.

---

**Figure 2A:**
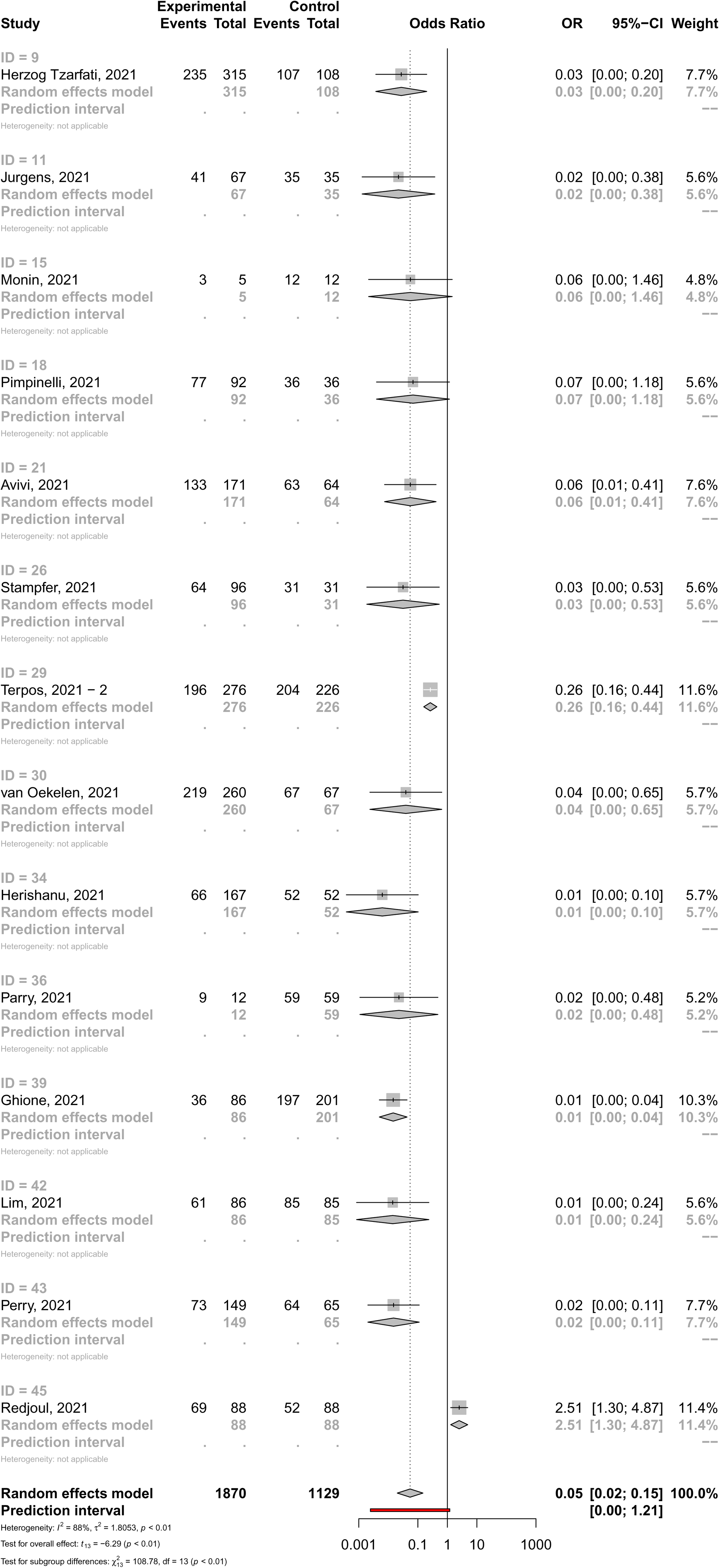
Odds ratio for achieving seropositivity in patients with haematological malignancy versus healthy control group following two doses of COVID-19 vaccine.

**Figure 2B:**
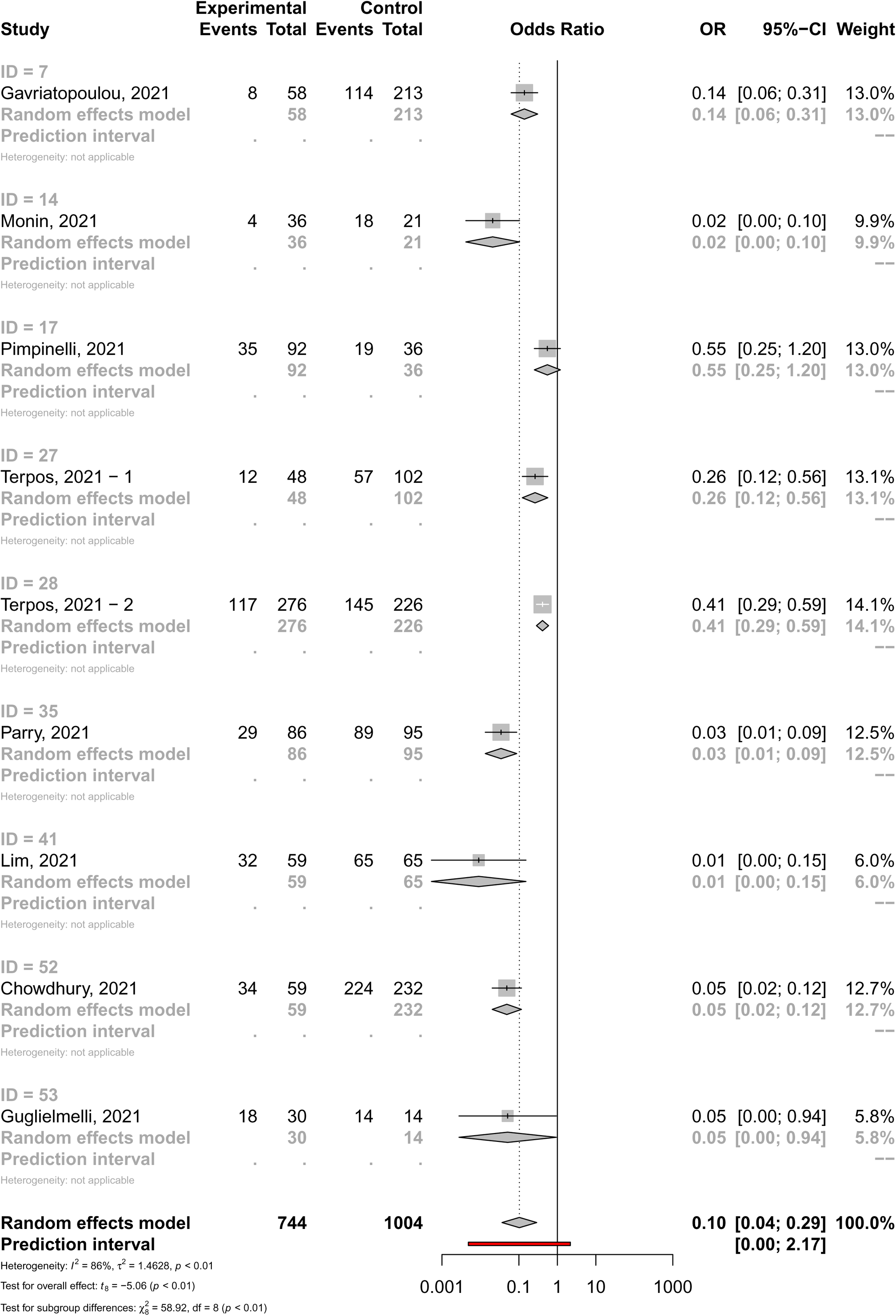
Odds ratio of achieving seropositivity in patients with haematological malignancy versus healthy control group following one dose of COVID-19 vaccine.

**Figure 3A:**
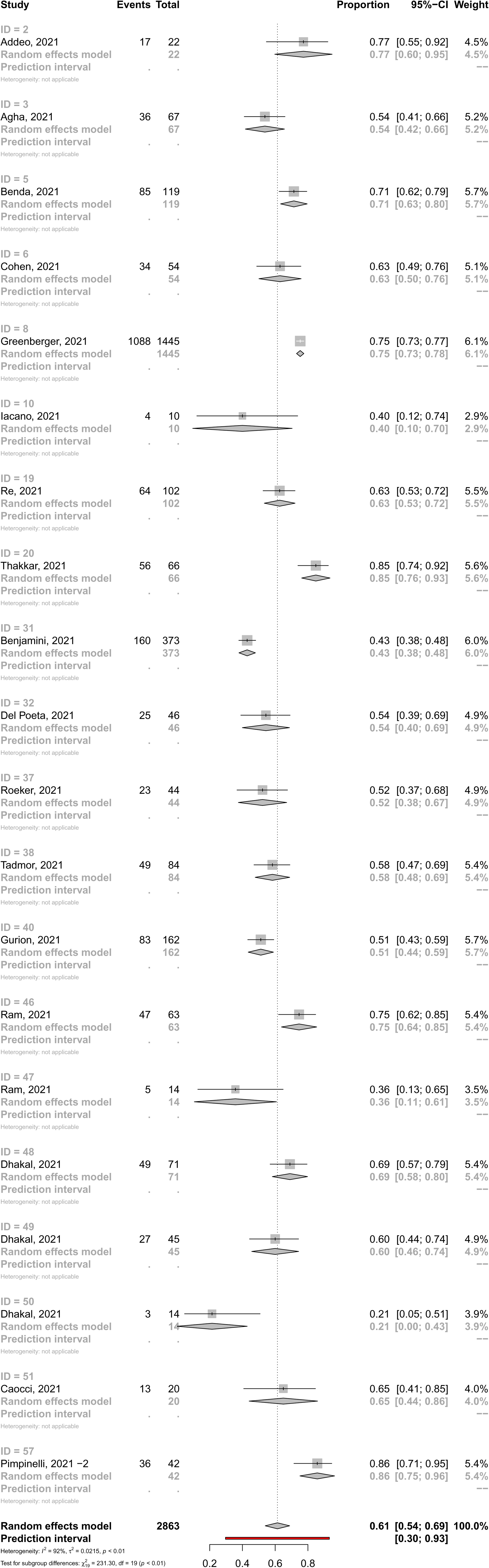
Pooled rates of seropositivity in single arm studies involving patients with haematological malignancy following two doses of COVID-19 vaccine.

**Figure 3B:**
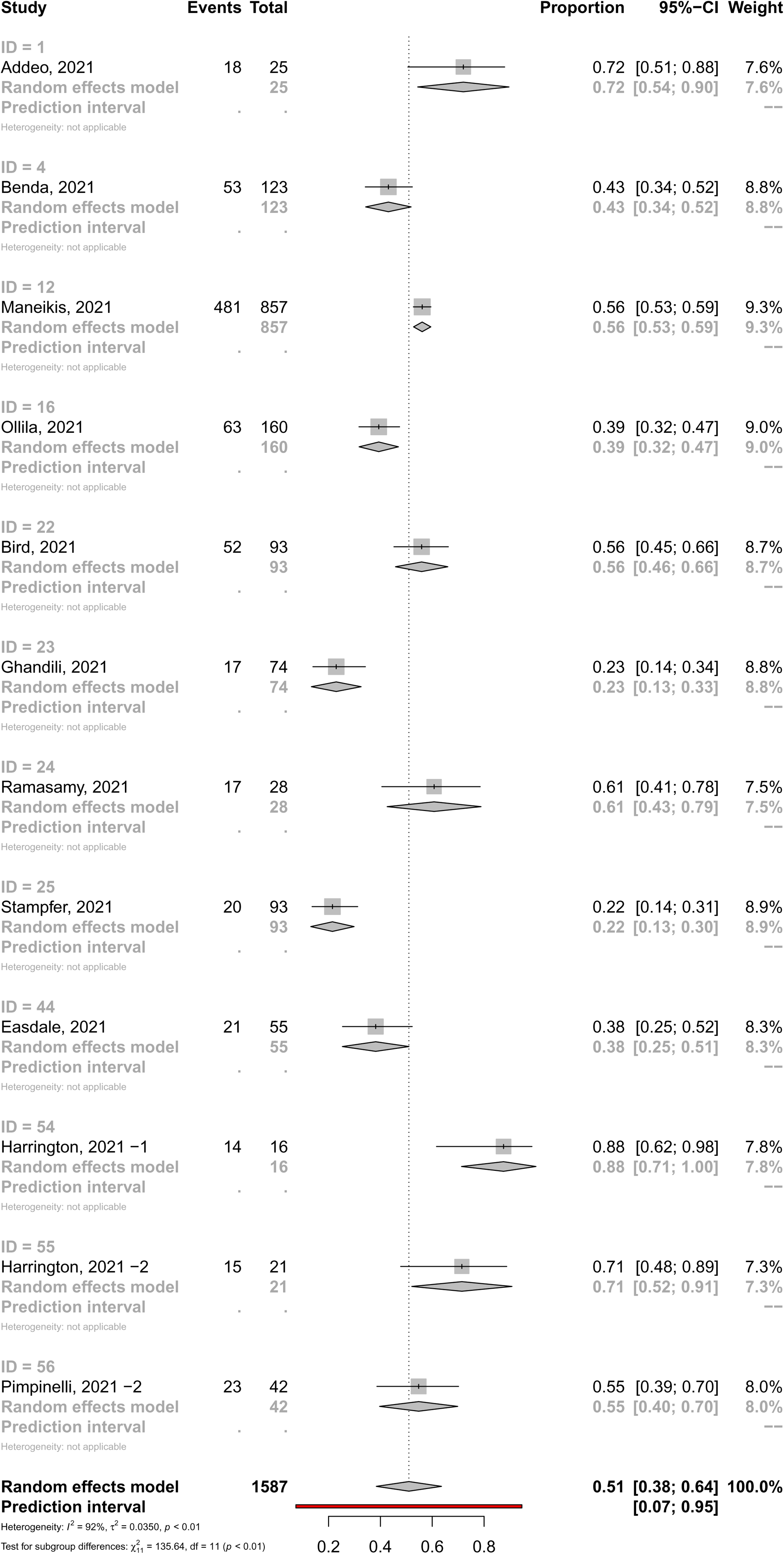
Pooled rates of seropositivity in single arm studies involving patients with haematological malignancy following one dose of COVID-19 vaccine.

## Notes

### Clinical Protocols

https://www.crd.york.ac.uk/prospero/display_record.php?RecordID=276851

### Funding Statement

This study did not receive funding

### Author Declarations

This study is a systematic review and meta-analysis of published studies reporting clinical data.

