## Supplementary data for "Immunogenicity of COVID-19 vaccines in patients with haematological malignancy: A systematic review and meta-analysis"

| Study (author, year) | Selection | | | | Comparability | Outcome | | | Rating |
|---|---|---|---|---|---|---|---|---|---|
| | Representativeness of the exposed cohort | Selection of the non-exposed cohort | Ascertainment of exposure (refers to diagnosis of HM) | Demonstration that outcome of interest was not present at start of study | Comparability of cohorts on the basis of the design or analysis | Assessment of outcome | Was follow-up long enough for outcomes to occur? | Adequacy of follow up of cohorts (% follow-up rate) | |
| Addeo et. al. 2021 | ✵ | NA | ✵ | ✵ | NA | ✵ | ✵ | ✵ | Good (A) |
| Agha et. al. 2021 | ✵ | NA | | | NA | ✵ | ✵ | ✵ | Poor (A) |
| Avivi et. al. 2021 | ✵ | ✵ | ✵ | ✵ | | ✵ | ✵ | ✵ | Poor |
| Benda et. al. 2021 | ✵ | NA | ✵ | | NA | ✵ | ✵ | ✵ | Fair (A) |
| Benjamini et. al. 2021 | ✵ | NA | ✵ | | NA | ✵ | ✵ | ✵ | Fair (A) |
| Bird et. al. 2021 | ✵ | NA | ✵ | | NA | ✵ | | ✵ | Fair (A) |
| Caocci et. al. 2021 | | NA | | | NA | ✵ | ✵ | ✵ | Poor (A) |
| Cohen et. al. 2021 | | NA | ✵ | | NA | ✵ | ✵ | ✵ | Poor (A) |
| Del Poeta et. al. 2021 | | NA | ✵ | | NA | ✵ | ✵ | ✵ | Poor (A) |
| Dhakal et. al. 2021 | | NA | ✵ | | NA | ✵ | | ✵ | Poor (A) |
| Easdale et. al. 2021 | | NA | ✵ | | NA | ✵ | ✵ | ✵ | Poor (A) |
| Gavriatopoulou et. al. 2021 | ✵ | ✵ | ✵ | | ✵ | ✵ | | ✵ | Good |
| Ghandili et. al. 2021 | ✵ | NA | ✵ | ✵ | NA | ✵ | | ✵ | Good (A) |

| | | | | | | | | | |
|---|---|---|---|---|---|---|---|---|---|
| Ghione et. al. 2021 | ✹ | ✹ | ✹ | ✹ | | ✹ | | ✹ | Poor |
| Greenberger et. al. 2021 | ✹ | NA | | ✹ | NA | ✹ | ✹ | ✹ | Fair (A) |
| Guglielmelli et. al. 2021 | ✹ | | | ✹ | | | | ✹ | Poor |
| Gurion et. al. 2021 | ✹ | NA | ✹ | | NA | ✹ | | ✹ | Fair (A) |
| Harrington et. al. 2021 (BrJH) | ✹ | NA | ✹ | | NA | ✹ | | ✹ | Fair (A) |
| Harrington et. al. 2021 (Leuk) | ✹ | NA | ✹ | | NA | ✹ | | ✹ | Fair (A) |
| Herishanu et. al. 2021 | ✹ | | ✹ | ✹ | ✹ | ✹ | ✹ | ✹ | Good |
| Herzog Tzarfati et. al. 2021 | ✹ | | ✹ | | ✹✹ | ✹ | ✹ | ✹ | Fair |
| Iacono et. al. 2021 | | ✹ | ✹ | ✹ | | ✹ | | | Poor |
| Jurgens et. al. 2021 | ✹ | ✹ | ✹ | | | ✹ | ✹ | ✹ | Poor |
| Lim et. al. 2021 | ✹ | ✹ | ✹ | ✹ . | | ✹ | | | Poor |
| Malard et. al. 2021 | ✹ | | ✹ | ✹ | | ✹ | | ✹ | Poor |
| Manekis et. al. 2021 | ✹ | ✹ | ✹ | ✹ | ✹ | ✹ | | | Poor |
| Monin et. al. 2021 | ✹ | ✹ | ✹ | ✹ | | ✹ | ✹ | | Poor |
| Ollila et. al. 2021 | | NA | ✹ | | NA | ✹ | ✹ | ✹ | Poor (A) |

| | | | | | | | | | |
|---|---|---|---|---|---|---|---|---|---|
| Parry et. al. 2021 | | ✹ | | | | ✹ | ✹ | | Poor |
| Perry et. al. 2021 | ✹ | | ✹ | ✹ | ✹ | ✹ | | ✹ | Good |
| Pimpinelli et. al. 2021 (PV/ET) | ✹ | NA | ✹ | ✹ | NA | ✹ | | ✹ | Good (A) |
| Pimpinelli et. al. 2021 (MM/MPN) | ✹ | | ✹ | ✹ | | ✹ | | ✹ | Poor |
| Ram et. al. 2021 | | NA | ✹ | ✹ | NA | ✹ | | ✹ | Fair (A) |
| Ramasamy et. al. 2021 | ✹ | NA | | | NA | ✹ | | | Poor (A) |
| Re et. al. 2021 | | NA | ✹ | | NA | | | ✹ | Poor (A) |
| Redjoul et.al. 2021 | ✹ | NA | ✹ | | NA | ✹ | ✹ | ✹ | Fair (A) |
| Roeker et. al. 2021 | ✹ | NA | ✹ | | NA | ✹ | ✹ | ✹ | Fair (A) |
| Stampfer et. al. 2021 | ✹ | ✹ | ✹ | | ✹ | ✹ | | ✹ | Good |
| Strickland et. al. 2021 | ✹ | ✹ | ✹ | ✹ | ✹ | ✹ | | ✹ | Good |
| Tadmor et. al. 2021 | ✹ | NA | ✹ | | NA | ✹ | ✹ | ✹ | Fair (A) |
| Terpos et. al. 2021 (Blood) | ✹ | ✹ | ✹ | ✹ | ✹ | ✹ | | ✹ | Good |
| Terpos et. al. 2021 (BCJ) | ✹ | ✹ | ✹ | | ✹ | ✹ | | ✹ | Good |
| Thakkar et. al. 2021 | ✹ | | ✹ | ✹ | | ✹ | ✹ | ✹ | Poor |
| Van Oekelen et. al. 2021 | ✹ | ✹ | ✹ | | ✹ | ✹ | ✹ | ✹ | Good |

Note: A study can be awarded a maximum of one star (✸) for each numbered item within the selection and outcome domains. A maximum of two stars can be given for comparability domain. A good quality study would score 3 or 4 stars in selection domain, 1 or 2 stars in comparability domain, and 2 or 3 stars in outcome domain. A fair quality study would require 2 stars in selection domain, 1 or 2 stars in comparability domain, and 2 or 3 stars in outcome domain. On the other hand, a study will be regarded as poor quality if it reflects 0 or 1 star in selection domain, or 0 star in comparability domain, or 0 or 1 star in outcome domain. For studies without a control group, item 2 in selection domain and comparability domain are not applicable. Therefore, the rating system proposed by Sharmin et al was adapted accordingly; a good quality study would score 3 stars in selection domain, and 2 or 3 stars in outcome domain. A fair quality study would require 2 stars in selection domain, and 2 or 3 stars in outcome domain. On the other hand, a study will be regarded as poor quality if it reflects 0 or 1 star in selection domain, or 0 or 1 star in outcome domain. In this review, a good or fair quality study is considered of having low risk of bias while a poor quality study is regarded as high risk of bias.

HM: haematological malignancy; PV: Polycythaemia vera; ET: Essential thrombocytosis; MM: multiple myeloma; MPN: myeloproliferative neoplasm; BCJ: Blood Cancer Journal; NA: not applicable; A (adapted)

**Supplementary Table 1: Summary of risk of bias/quality assessment of included studies**

| Study | Type /Location | Study population/ Comparator | Number of participants (analysed) | Age Median | Male/ Female | Vaccine type | Analysis | Seropositivity | Rate of positive neutralising antibody/ cellular response | Adverse events |
|---|---|---|---|---|---|---|---|---|---|---|
| Avivi et. al. | Single centre Prospective cohort study<br><br>Israel | Myeloma<br><br>Healthy volunteers | 171 patients<br><br>64 controls | 70 years (38-94) | 96 Male<br>75 Female | BNT162b2 | SARS-CoV-2 S IgG Roche ≥ 0.8 UI/ml = positive | 14-21 days post second dose: 133/171 (78%) vs. 63/64 (98%) controls | Not reported | At least one adverse event: 90/161 (53%) vs. 29/53 (55%) controls |
| Bird et. al. | Single centre retrospective cohort study<br><br>United Kingdom | Myeloma | 93 patients | 67 years (47-87) | 55 Male<br>38 Female | BNT162b2<br>ChAdOx1<br><br>52%<br>48% | SARS-CoV-2 S IgG Ortho clinical ≥1 signal/cut-off = positive | ≥ 21 days post first dose: 52/93 (56%) | Not reported | Not reported |
| Ghandili et. al. | Single centre prospective cohort study<br><br>Germany | Myeloma | 82 patients | 68 years (40-85) | 49 Male<br>33 Female | BNT162b2<br>ChAdOx1<br><br>77%<br>23% | SARS-CoV-2 S IgG DiaSorin ≥ 34 AU/ml = positive | 21 days post first dose: 17/74 (23%) | Not reported | Not reported |
| Ramasamy et. al. | Multicentre web-based prospective cohort study United | Myeloma | 105 patients -28 patients sampled | 63 years | 67 Male<br>42 Female | BNT162b2<br>ChAdOx1<br><br>42%<br>58% | SARS-CoV-2 S IgG Abbott COI ≥ 50 = | >21 days post first dose: 17/28 (61%) | Not reported | Not reported |

| | | | | | | | | | | |
|---|---|---|---|---|---|---|---|---|---|---|
| | Kingdom | | | | | | positive | | | |
| Stampfer et. al. | Single centre prospective cohort study<br><br>United States | Myeloma<br><br><br>Healthy controls<br>Pre-COVID-19 controls | 103 patients<br><br><br>31 controls<br><br>34 controls | 68 years (35-88) | 61 Male<br>42 Female | BNT162b2<br>mRNA-1273<br><br>50%/50% | SARS-CoV-2 S IgG 'in house' 50-250 IU/ml = positive (partial response)<br><br>>250 IU/ml = clinically relevant response | 14-21 days post first dose:<br>20/93 (22%)<br><br>14-21 days post second dose:<br>64/96 (67%)<br>vs.<br>31/31 (100%) controls<br><br>>250 IU/ml<br>14-21 days post first dose:<br>2/93 (2%)<br><br>14-21 days post second dose:<br>43/93 (46%)<br>vs.<br>29/31 (94%) controls | Not reported | Not reported |
| Terpos et. al. | Single centre prospective cohort study<br><br>Greece | Myeloma<br><br><br>Matched Controls | 48 patients<br><br><br>102 controls | 83 years (59-92) | 29 Male<br>19 Female | BNT162b2 | SARS-CoV-2 neutralising Ab Genscript<br>≥ 30% = positive<br>≥ 50% = clinically relevant | ≥ 30%<br>22 days post first dose:<br>12/48 (25%)<br>vs.<br>57/102 (55%) controls | ≥ 50%<br>22 days post first dose:<br>4/48 (8%)<br>vs.<br>21/102 (20%) controls | Not reported |

| | | | | | | | | | | |
|---|---|---|---|---|---|---|---|---|---|---|
| Terpos et. al. | Single centre prospective cohort study<br><br>Greece | Myeloma<br><br><br><br><br>Matched Controls | 276 patients<br>-77% myeloma<br>-14% sMM<br>-9% MGUS<br><br>226 controls | 74 years (62-80) | 151 Male<br>125 Female | BNT162b2<br>ChAdOx1<br><br>78%<br>22% | SARS-CoV-2 neutralising Ab Genscript<br>≥ 30% = positive<br>≥ 50% = clinically relevant | Day 22 post first dose:<br>117/276 (42%)<br>vs.<br>145/226 (64%) controls<br><br>Day 50 post first dose:<br>196/276 (71%)<br>vs.<br>204/226 (90%) controls | Day 22 post first dose:<br>55/276 (20%)<br>vs.<br>73/226 (32%) controls<br><br>Day 50 post first dose:<br>158/276 (57%)<br>vs.<br>183/226 (81%) controls | First dose BNT162b2<br>71/215 (33%) local reaction<br><br>28/215 (13%) systemic reaction<br><br>ChAdOx1<br>20/61 (33%) local reaction<br><br>Second dose BNT162b2<br>68/215 (32%) local reaction<br><br>45/215 (21%) systemic reaction |
| Van Oekelen et. al. | Single centre prospective and retrospective cohort study<br><br>United States | Myeloma<br><br><br><br><br><br>Matched control health care workers | 320 patients<br>-260 sampled<br><br><br><br><br>67 controls | 68 years (38-93) | 185 Male<br>135 Female | BNT162b2<br>mRNA-1273<br>unknown<br><br>69%<br>27%<br>4% | SARS-CoV-2 S IgG<br>'in house'<br>≥ 5 AU/ml = positive | 51 days post second dose:<br>219/260 (84%)<br>vs.<br>67/67 (100%) controls | Not reported | Not reported |

| | | | | | | | | | | |
|---|---|---|---|---|---|---|---|---|---|---|
| **Subset of myeloma patients from other studies** | | | | | | | | | | |
| Agha et. al. | Single centre retrospective cohort study<br><br>United States | Haematology Myeloma subset | 67 patients | 71 years (IQR 65-77) | 35 Male<br>32 Female | BNT162b2<br>mRNA-1273<br><br>51%/42%<br>7% unknown | SARS-CoV-2 S IgG Beckman Coulter ≥1.00 = positive | 21 days post second dose: 19/29 (66%) | Not reported | Not reported |
| Benda et. al. | Single centre prospective cohort study | Haematology Myeloma subset<br><br><br>Solid tumour | 123 patients -34% myeloma<br><br><br>136 patients | Not reported in haem | Not reported in haem | BNT162b2 | SARS-CoV-2 S IgG Roche ≥ 0.8 IU/ml = positive | 28 days post second dose: 25/34 (74%) | Not reported | Not reported for haem |
| Cohen et. al. | Single centre retrospective study<br>Israel | Haematology Myeloma subset | 54 patients -37% myeloma | 69 years (IQR 61-77) | 32 Male<br>22 Female | BNT162b2 | SARS-CoV-2 S IgG Roche ≥ 0.8 IU/ml = positive | 14-21 days post second dose: 16/20 (80%) | Not reported | Not reported |
| Greenberger et. al. | Multicentre prospective cohort study<br><br>United States | Haematology Myeloma subset | 1445 patients -15% myeloma | 68 years (16-110) | 574 Male<br>871 Female | BNT162b2<br>mRNA-1273<br><br>45%<br>55% | SARS-CoV-2 S IgG Roche ≥ 0.8 IU/ml = positive | >14 days post second dose: 204/213 (96%) | Not reported | Not reported |
| Herzog Tzarfati et. al. | Single centre prospective cohort study<br><br><br>Israel | Haematology Myeloma subset<br><br><br>Matched Healthy control | 315 patients Myeloma -16%<br><br><br>108 controls | 70 years (IQR 61-77) | 223 Male<br>200 Female | BNT162b2 | SARS-CoV-2 S IgG DiaSorin ≥ 12 AU/ml = positive | 30-60 days post second dose: 40/53 (76%) vs. 107/108 (99%) control | Not reported | Not reported |

| | | | | | | | | | | |
|---|---|---|---|---|---|---|---|---|---|---|
| Ollila et. al. | Single centre retrospective cohort study<br><br>United States | Haematology Myeloma subset | 160 patients<br>-15% plasma cell | 72 years (65-79) | 86 Male<br>74 Female | BNT162b2<br>mRNA-1273<br>Ad26 | SARS-CoV-2 S IgG Abbott Signal/cutoff ratio ≥1.4 = positive | 56 days post first dose:<br>14/24 (58%) myeloma | Not reported | Not reported |
| Monin et. al. | Multicentre prospective cohort study<br><br>United Kingdom | Haematology Myeloma subset<br><br>Health care workers controls | 56 patients<br>-68% B cell malignancy<br>-9% T cell malignancy<br>-18% myeloid/ acute leukaemia<br>-5% others<br><br>54 controls | 73 years (IQR 65-80) | Not extractable for haem | BNT162b2 | SARS-CoV-2 Spike IgG ≥ 70 EC50 = positive<br><br>SARS-CoV-2 specific T cells secreting IFN-gamma and/or IL2 >7 cytokine-secreting cells per $10^6$ PBMC = positive | 21 days post first dose:<br>3/9 (33%)<br>vs.<br>32/34 (94%) Controls<br><br>35 days post first dose:<br>1/7 (14%)<br>vs.<br>18/21 (86%) controls<br><br>35 days post first dose (with second dose):<br>1/1 (100%)<br>vs.<br>12/12 controls (100%) | Cellular response<br>21 days post first dose:<br>2/3 (66%)<br>vs.<br>14/17 (82%) controls<br><br>35 days post first dose:<br>1/4 (25%)<br>vs.<br>9/13 (69%) controls<br><br>35 days post first dose (with second dose):<br>1/1 (100%)<br>vs.<br>3/3 (100%) controls | Not extractable for haem |
| Pimpinelli et. al. | Single centre prospective study<br><br>Italy | Haematology Myeloma subset<br><br>Older age (> 80 years) | 42 patients<br><br>36 controls | 73 years (47-78) | 23 Male<br>19 Female | BNT162b2 | SARS-CoV-2 S IgG DiaSorin ≥ 15 AU/ml = positive | 21 days post first dose:<br>9/42 (21%) myeloma<br>vs.<br>19/36 (53%) | Not reported | Reported with different patient numbers |

| | | | | | | | | | | |
|---|---|---|---|---|---|---|---|---|---|---|
| | | control group | | | | | | controls<br><br>14 days post second dose: 33/42 (79%) myeloma vs. 36/36 (100%) controls<br><br>≥ 80 AU/ml 14 days post second dose 23/42 (55%) myeloma vs. 34/36 (97%) control | | |
| Re et. al. | Multicentre retrospective cohort study<br><br>France | Haematology Myeloma subset | 102 patients -22% myeloma | 76 years (33-93) | 67 Male<br>35 Female | BNT162b2<br>mRNA-1273<br><br>93%<br>7% | SARS-CoV-2 S IgG<br>Range of commercial platforms | 21-28 days post second dose<br>17/23 (74%) | Not reported | Not reported |

**Supplementary Table 2: Summary of study characteristics and outcomes for patients with myeloma**

| Study | Type /Location | Study population/ Comparator | Number of participants (analysed) | Age Median | Male/ Female | Vaccine type | Analysis | Seropositivity | Rate of positive neutralising antibody/ cellular response | Adverse events |
|---|---|---|---|---|---|---|---|---|---|---|
| Benjamini et. al. | Multicentre prospective cohort study<br><br>Israel | CLL patients | 373 patients | 70 years (40-89) | 222 Male<br>151 Female | BNT162b2 | SARS-CoV-2 S IgG DiaSorin ≥ 15 AU/ml = positive Abbott >50 U/ml = positive 'in house' >1.1 = positive | 14-21 days pot second dose: 160/373 (43%) | Not reported | At least 1 adverse event: 151/331 (47%) |
| Del Poeta et. al. | Single centre prospective cohort study<br><br>Italy | CLL patients | 46 patients | Not reported | 29 Male<br>17 Female | BNT162b2 | SARS-CoV-2 S IgG Maglumi ≥1.1 = positive | 14-21 days post second dose: 25/46 (54%) | Not reported | Not reported |
| Herishanu et. al. | Single centre prospective cohort study<br><br>Israel | CLL patients<br><br>Control -age, sex matched | 167 patients<br><br>52 controls | 71 years (63-76) | 112 Male<br>55 Female | BNT162b2 | SARS-CoV-2 S IgG Roche ≥ 0.8 IU/ml = positive | 14-21 days post second dose: 66/167 (40%) patients vs. 52/52 (100%) controls | Not reported | First dose 52/167 (31%) local reaction<br><br>21/167 (13%) Systemic reaction Second dose 56/167 (34%) local |

| | | | | | | | | | | |
|---|---|---|---|---|---|---|---|---|---|---|
| | | | | | | | | | | reaction<br><br>21/167 (23%) Systemic reaction |
| Parry et. al. | Single centre prospective cohort study<br><br>United Kingdom | CLL patients<br><br>Healthy age matched controls | 299 patients<br><br>93 controls | 69 years (IQR 63-74) | 159 Male<br>140 Female | BNT162b2<br>ChAxOd1<br><br>52%/48% | SARS-CoV-2 S IgG Roche ≥ 0.8 IU/ml = positive<br><br>Dried blood sampling Roche ratio ≥ 1.0 = positive | 43 days post first dose:<br>Serum<br>29/86 (34%)<br>vs.<br>89/95 (94%)<br>Control<br><br>Dried blood<br>63/267 (24%)<br>vs.<br>66/93 (71%)<br>control<br><br>18 days post second dose:<br>Serum<br>9/12 (75%)<br>vs.<br>59/59 (100%)<br>Controls<br><br>Dried blood<br>39/55 (71%)<br>vs.<br>36/37 (97%)<br>controls | Not reported | Not reported |
| Roeker et. al. | Single centre retrospective cohort study | CLL patients | 44 patients | 71 years (37-89) | 23 Male<br>21 Female | BNT162b2<br>mRNA-1273 | SARS-CoV-2 S IgG | 21 days post second dose:<br>23/44 (52%) | Not reported | Not reported |

| | | | | | | | | | | |
|---|---|---|---|---|---|---|---|---|---|---|
| | United States | | | | | 57%<br>43% | DiaSorin<br>≥ 15 AU/ml<br>= positive | | | |
| Tadmor et. al. | Multicentre prospective observation study<br><br>Israel | CLL patients | 84 patients | 69 years (44-87) | 53 Male<br>29 Female | BNT162b2 | SARS-CoV-2 S IgG<br>Abbott<br>≥ 50 U/ml = positive<br><br>SARS-CoV-2 RBD IgG<br>>1.1 = positive | 22 days post second dose: 49/84 (58%) | Not reported | Not reported |
| **Subset of CLL patients from other haematology studies** | | | | | | | | | | |
| Agha et. al. | Single centre retrospective cohort study<br><br>United States | Haematology CLL subset | 67 patients<br>-19% CLL | 71 years (IQR 65-77) | 35 Male<br>32 Female | BNT162b2<br>mRNA-1273<br><br>51%<br>42%<br>7% unknown | SARS-CoV-2 S IgG<br>Beckman Coulter<br>≥1.00 = positive | 21 days post second dose: 3/13 (23%) | Not reported | Not reported |
| Greenberger et. al. | Multicentre prospective cohort study<br><br>United States | Haematology CLL subset | 1445 patients<br>-45% CLL | 68 years (16-110) | 574 Male<br>871 Female | BNT162b2<br>mRNA-1273<br><br>45%<br>55% | SARS-CoV-2 S IgG<br>Roche<br>≥ 0.8 IU/ml = positive | >14 days post second dose: 417/650 (64%) | Not reported | Not reported |
| Herzog Tzarfati et. al. | Single centre prospective cohort study<br><br>Israel | Haematology CLL subset<br><br><br>Matched | 315 patients<br>-11% CLL<br><br><br>108 controls | 70 years (IQR 61-77) | 223 Male<br>200 Female | BNT162b2 | SARS-CoV-2 S IgG<br>DiaSorin<br>≥ 12 AU/ml | 30-60 days post second dose:<br>16/34 (46%) vs. | Not reported | Not reported |

| | | | | | | | | | | |
|---|---|---|---|---|---|---|---|---|---|---|
| | | Healthy control | | | | | = positive | 107/108 (99%) control | | |
| Jurgens et. al. | Single centre prospective cohort study<br><br>United States | Haematology CLL subset<br><br><br>Control health care workers | 67 patients<br>-31% CLL<br><br><br>35 controls | 71 years (24-90) | 36 Male<br>31 Female | BNT162b2<br>mRNA-1273<br><br>46%<br>54% | SARS-CoV-2 S IgG 'in house' OD450 ≥ 3 = positive | 21 days post second dose:<br>12/21 (57%)<br>vs.<br>35/35 (100%) controls | Not reported | Not reported |
| Monin et. al. | Multicentre prospective cohort study<br><br>United Kingdom | Haematology CLL subset<br><br><br><br><br><br><br><br><br><br><br><br>Health care workers controls | 56 patients<br>-68% B cell malignancy<br>-9% T cell malignancy<br>-18% myeloid/ acute leukaemia<br>-5% others<br><br><br><br><br>54 controls | 73 years (IQR 65-80) | Not extractable for haem | BNT162b2 | SARS-CoV-2 Spike IgG ≥ 70 EC50 = positive<br><br><br>SARS-CoV-2 specific T cells secreting IFN-gamma and/or IL2 >7 cytokine-secreting cells per $10^6$ PBMC = positive | 21 days post first dose:<br>1/6 (17%)<br>vs.<br>32/34 (94%) Controls<br><br>35 days post first dose:<br>0/6 (0%)<br>vs.<br>18/21 (86%) controls<br><br>35 days post first dose (with second dose):<br>1/2 (50%)<br>vs.<br>12/12 (100%) controls | Cellular response 21 days post first dose:<br>2/5 (40%)<br>vs.<br>14/17 (82%) controls<br><br>35 days post first dose:<br>1/4 (25%)<br>vs.<br>9/13 (69%) controls<br><br>35 days post first dose (with second dose):<br>1/1 (100%)<br>vs.<br>3/3 (100%) controls | Not extractable for haem |
| Ollila et. al. | Single centre retrospective cohort study | Haematology CLL subset | 160 patients<br>-12% CLL | 72 years (65-79) | 86 Male<br>74 Female | BNT162b2<br>mRNA-1273<br>Ad26 | SARS-CoV-2 S IgG Abbott | 56 days post first dose:<br>7/19 (37%) CLL | Not reported | Not reported |

| | | | | | | | |
|---|---|---|---|---|---|---|---|
| | United States | | | | | | Signal/cutoff ratio ≥1.4 = positive |

**Supplementary Table 3: Summary of study characteristics and outcomes for patients with chronic lymphocytic leukaemia**

| Study | Type /Location | Study population/ Comparator | Number of participants (analysed) | Age Median | Male/ Female | Vaccine type | Analysis | Seropositivity | Rate of positive neutralising antibody/ cellular response | Adverse events |
|---|---|---|---|---|---|---|---|---|---|---|
| Ghione et. al. | Single centre prospective cohort study<br><br>United States | B-cell lymphoma<br><br>Control age-care, healthcare workers | 86 patients<br><br><br>47 controls<br><br>154 controls | 70 years (35-91) | 45 Male<br>41 Female | BNT162b2<br>mRNA-1273<br>Ad26<br><br>47%<br>52%<br>1% | SARS-CoV-2 S IgG BioRad ≥1.0 = positive | 14-56 days post completion of vaccination: 36/86 (42%) patients vs. 43/47 (92%) age-care<br><br>154/154 (100%) healthcare | Not reported | Not reported |
| Gurion et. al. | Multicentre prospective cohort study<br><br>Israel | Lymphoma | 162 patients<br>-88% NHL<br>-12% HL | 65 years (52-73) | 89 Male<br>73 Female | BNT162b2 | SARS-CoV-2 S IgG Abbott ≥ 50 IU/ml = positive | 28 days post second dose: 83/162 (51%) | Not reported | Not reported |
| Lim et. al. | Multicentre prospective cohort study Interim analysis<br><br><br>United Kingdom | Lymphoma | 129 patients recruited<br>119 analysed<br>-66% indolent B-NHL<br>-29% aggressive B-NHL<br>-10% HL<br>-3% other | 69 years (IQR 57-74) | 81 Male<br>48 Female | BNT162b2<br>ChAdOx1 | SARS-CoV-2 S IgG Meso Scale Discovery >0.55 BAU/ml = positive<br><br>RBD IgG >0.73 BAU/ml = | 14 days post first dose: 32/59 (54%) patients vs. 65/65 (100%) controls<br><br>14-28 days post second dose: 61/86 (71%) | Not reported | Not reported |

| | | | | | | | | | | |
|---|---|---|---|---|---|---|---|---|---|---|
| | | Healthy control | 150 control | | | | positive | patients<br>vs.<br>85/85 (100%) controls | | |
| Perry et. al. | Single centre prospective cohort study<br><br>Israel | Lymphoma<br>-B cell NHL<br><br>Healthy control | 149 patients<br>-53% indolent NHL<br>-47% aggressive NHL<br><br>65 controls | 64 years (20-92) | 88 Male<br>61 Female | BNT162b2 | SARS-CoV-2 S IgG<br>Roche<br>≥ 0.8 IU/ml = positive | 14-21 days post second dose:<br>73/149 (49%)<br>vs.<br>64/65 (99%) controls<br><br>38/80 (48%) indolent NHL<br><br>34/69 (49%) aggressive NHL | Not reported | At least 1 adverse event:<br>60/118 (51%)<br><br>44/118 (37%) local AE<br><br>23/118 (20%) Systemic AE |
| **Subset of lymphoma patients from other haematology studies** | | | | | | | | | | |
| Agha et. al. | Single centre retrospective cohort study<br><br>United States | Haematology Lymphoma subset | 67 patients<br>-31% lymphoma | 71 years (IQR 65-77) | 35 Male<br>32 Female | BNT162b2<br>mRNA-1273<br><br>51%/42%<br>7% unknown | SARS-CoV-2 S IgG<br>Beckman Coulter<br>≥1.00 = positive | 21 days post second dose:<br>11/21 (52%) | Not reported | Not reported |
| Cohen et. al. | Single centre retrospective cohort study<br>Israel | Haematology Lymphoma subset | 54 patients<br>-61% lymphoma | 69 years (IQR 61-77) | 32 Male<br>22 Female | BNT162b2 | SARS-CoV-2 S IgG<br>Roche<br>≥ 0.8 IU/ml = positive | 14-21 days post second dose:<br>17/33 (52%) | Not reported | Not reported |
| Greenberger et. al. | Multicentre prospective | Haematology Lymphoma | 1445 patients | 68 years (16-110) | 574 Male<br>871 Female | BNT162b2<br>mRNA-1273 | SARS-CoV-2 | >14 days post second dose: | Not reported | Not reported |

| | | | | | | | | | | |
|---|---|---|---|---|---|---|---|---|---|---|
| | cohort study<br><br>United States | subset | -25% NHL<br>-5% HL | | | <br>45%<br>55% | S IgG<br>Roche<br>≥ 0.8 IU/ml<br>= positive | 266/363<br>(73%)<br>NHL<br><br>64/65<br>(98%)<br>HL | | |
| Herzog Tzarfati et. al. | Single centre prospective cohort study<br><br>Israel | Haematology<br><br><br><br><br><br><br><br><br><br>Matched Healthy control | 315 patient<br>-16% aggressive NHL<br>-13% indolent NHL<br>-5% HL<br><br><br><br><br>108 controls | 70 years (IQR 61-77) | 223 Male<br>200 Female | BNT162b2 | SARS-CoV-2 S IgG DiaSorin<br>≥ 12 AU/ml<br>= positive | 30-60 days post second dose:<br>36/51 (71%) aggressive NHL<br><br>24/40 (60%) Indolent NHL<br><br>15/17 (94%) HL<br>vs.<br>107/108 (99%) control | Not reported | Not reported |
| Jurgens et. al. | Single centre prospective cohort study<br><br>United States | Haematology<br><br><br><br>Control health care workers | 67 patients<br>-63% NHL<br>-6% HL<br><br>35 controls | 71 years (24-90) | 36 Male<br>31 Female | BNT162b2<br>mRNA-1273<br><br>46%/54% | SARS-CoV-2 S IgG<br>'in house'<br>OD450 ≥ 3<br>= positive | 21 days post second dose:<br>25/42 (60%) NHL<br><br>4/4 (100%) HL<br>vs.<br>35/35 (100%) controls | Not reported | Not reported |
| Monin et. al. | Multicentre prospective cohort study<br><br>United Kingdom | Haematology<br>Lymphoma subset | 56 patients<br>-68% B cell malignancy<br>-9% T cell malignancy<br>-18% myeloid/ | 73 years (IQR 65-80) | Not extractable for haem | BNT162b2 | SARS-CoV-2 Spike IgG<br>≥ 70 EC50<br>= positive | 21 days post first dose:<br>2/15 (13%)<br>vs.<br>32/34 (94%) Controls | Cellular response<br>21 days post first dose:<br>3/3 (100%)<br>vs.<br>14/17 (82%) | Not extractable for haem |

| | | | | | | | | | |
|---|---|---|---|---|---|---|---|---|---|
| | | Health care workers controls | acute leukaemia<br>-5% others<br><br>54 controls | | | | SARS-CoV-2 specific T cells secreting IFN-gamma and/or IL2 >7 cytokine-secreting cells per $10^6$ PBMC = positive | 35 days post first dose:<br>1/10 (10%)<br>vs.<br>18/21 (86%) controls<br><br>35 days post first dose (with second dose):<br>0/1 (0%)<br>vs.<br>12/12 (100%) controls | controls<br><br>35 days post first dose:<br>1/3 (33%)<br>vs.<br>9/13 (69%) controls<br><br>35 days post first dose (with second dose):<br>0/1 (0%) vs.<br>3/3 (100%) controls | |
| Ollila et. al. | Single centre retrospective cohort study<br><br>United States | Haematology Lymphoma subset | 160 patients<br>-36% aggressive lymphoma<br>-21% indolent lymphoma<br>- 9% other lymphoma | 72 years (65-79) | 86 Male<br>74 Female | BNT162b2<br>mRNA-1273<br>Ad26 | SARS-CoV-2 S IgG Abbott Signal/cutoff ratio ≥1.4 = positive | 56 days post first dose:<br>27/107 (25%) Total lymphoma<br><br>9/58 (16%) Aggressive lymphoma<br><br>12/34 (35%) indolent lymphoma<br><br>6/15 (40%) Other lymphoma | Not reported | Not reported |

**Supplementary Table 4: Summary of study characteristics and outcomes for patients with lymphoma**

| Study | Type /Location | Study population/ Comparator | Number of participants (analysed) | Age Median | Male/ Female | Vaccine type | Analysis | Seropositivity | Rate of positive neutralising antibody/ cellular response | Adverse events |
|---|---|---|---|---|---|---|---|---|---|---|
| Easdale et. al. | Single centre retrospective cohort study<br><br>United Kingdom | Allogeneic HCT<br>>3 months | 55 patients | 50 years (18-73) | 34 Male<br>21 Female | BNT162b2<br>ChAdOx1<br><br>38%<br>62% | SARS-CoV-2<br>S IgG<br>Ortho clinical<br>≥1 signal/cut-off = positive | 42 days post first dose:<br>21/55 (38%) | Not reported | Not reported |
| Redjoul et. al. | Single centre retrospective cohort study<br><br>France | Allogeneic HCT | 88 patients | Not reported | 47 Male<br>41 Female | BNT162b2 | SARS-CoV-2<br>S IgG<br>Abbott<br>>21 AU/ml = positive<br><br>>4160 AU/ml = neutralisati on | 28 days post second dose:<br>69/88 (78%)<br><br>>4160 AU/ml<br>28 days post second dose:<br>52/88 (59%) | Not reported | Not reported |
| Ram et. al. | Single centre prospective cohort study<br><br>Israel | Allogeneic HCT and CAR-T<br>>3 months | 80 patients<br>-83% alloHCT<br>-17% CAR-T | 65 years (23-83) | 44 Male<br>37 Female | BNT162b2 | SARS-CoV-2 S IgG<br>Roche<br>≥ 0.8 U/ml = positive<br><br>SARS-CoV-2 specific cells | 7 to 14 days post second dose:<br>47/63 (75%) alloHCT<br><br>5/14 (36%) CAR-T | Cellular<br>7 to 14 days post second dose:<br><br>7/37 (19%) alloHCT<br><br>6/12 (50%) CAR-T | At least 1 adverse event:<br>First dose 11/80 (14%)<br><br>3/80 (4%) GvHD<br><br>Second dose 18/74 (24%) |

| | | | | | | | | | | |
|---|---|---|---|---|---|---|---|---|---|---|
| | | | | | | | ELISPOT, (IFN, IL2) 4 spots/well = positive | | | 3/74 (4%) GvHD |
| Dhakal et. al. | Single centre retrospective cohort study<br><br>United States | Autologous Allogeneic HCT CAR-T | 130 patients -45 autoHCT -71 alloHCT -14 CAR-T | autoHCT 65 years (45-75) alloHCT 64 years (25-77)<br><br>Age not specified for CAR-T | Not reported | BNT162b2 mRNA-1273 Ad26<br><br>59% 36% 5% | SARS-CoV-2 S IgG EUROIMMUN ≥1.1 signal/cut-off = positive | 14 days post completion of vaccination: 27/45 (60%) autoHCT 49/71 (38%) alloHCT 3/14 (21%) CAR-T | Not reported | Not reported |
| **Subset of HCT patients from other haematology studies** | | | | | | | | | | |
| Herzog Tzarfati et. al. | Single centre prospective cohort study<br><br>Israel | Haematology Autologous HCT subset<br><br>Matched Healthy control | 315 patients<br><br>108 controls | 70 years (IQR 61-77) | 223 Male 200 Female | BNT162b2 | SARS-CoV-2 S IgG DiaSorin ≥ 12 AU/ml = positive | 30-60 days post second dose: 17/21 (81%) autoHCT vs. 107/108 (99%) control | Not reported | Not reported |
| Greenberger et. al. | Multicentre prospective cohort study<br><br>United States | Haematology HCT and CAR-T subset | 1445 patients | 68 years (16-110) | 574 Male 871 Female | BNT162b2 mRNA-1273<br><br>45% 55% | SARS-CoV-2 S IgG Roche ≥ 0.8 IU/ml = positive | >14 days post second dose: 65/73 (89%) HCT<br><br>5/12 (42%) CAR-T | Not reported | Not reported |

**Supplementary Table 5: Summary of study characteristics and outcomes for patients following haematopoietic stem cell transplant and cellular therapy**

| Study | Type /Location | Study population/ Comparator | Number of participants (analysed) | Age Median | Male/ Female | Vaccine type | Analysis | Seropositivity | Rate of positive neutralising antibody/ cellular response | Adverse events |
|---|---|---|---|---|---|---|---|---|---|---|
| **Subset of acute leukaemia and myelodysplastic syndrome patients from other haematology studies** | | | | | | | | | | |
| Benda et. al. | Single centre prospective cohort study | Haematology AML subset<br><br>Solid tumour | 123 patients -28% AML/MDS/ MPN<br><br>136 patients | Not reported in haem | Not reported in haem | BNT162b2 | SARS-CoV-2 S IgG Roche ≥ 0.8 IU/ml = positive | 28 days post second dose: 33/34 (97%) | Not reported | Not reported for haem |
| Greenberger et. al. | Multicentre prospective cohort study<br><br>United States | Haematology Acute leukaemia subset | 1445 patients -4% acute leukaemia | 68 years (16-110) | 574 Male 871 Female | BNT162b2 mRNA-1273<br><br>45%<br>55% | SARS-CoV-2 S IgG Roche ≥ 0.8 IU/ml = positive | >14 days post second dose: 46/51 (90%) | Not reported | Not reported |
| Herzog Tzarfati et. al. | Single centre prospective cohort study<br><br>Israel | Haematology Acute leukaemia MDS subset<br><br>Matched Healthy control | 315 patients -5% Acute leukemia -5% MDS<br><br>108 controls | 70 years (IQR 61-77) | 223 Male 200 Female | BNT162b2 | SARS-CoV-2 S IgG DiaSorin ≥ 12 AU/ml = positive | 30-60 days post second dose: 12/15 (80%) acute leukaemia<br><br>15/16 (94%) MDS vs. 107/108 (99%) control | Not reported | Not reported |
| Monin et. al. | Multicentre prospective cohort study | Haematology Acute leukaemia subset | 56 patients -18% myeloid, leukaemia | 73 years (IQR 65-80) | Not extractable for haem | BNT162b2 | SARS-CoV-2 Spike IgG ≥ 70 EC50 | 21 days post first dose: 0/3 (0%) vs. | Cellular response 21 days post first dose: | Not extractable for haem |

| | | | | | | | | | | |
|---|---|---|---|---|---|---|---|---|---|---|
| | United Kingdom | Health care workers controls | -5% others<br><br>54 controls | | | | = positive<br><br>SARS-CoV-2 specific T cells secreting IFN-gamma and/or IL2 >7 cytokine-secreting cells per $10^6$ PBMC = positive | 32/34 (94%) Controls<br><br>35 days post first dose:<br>0/3 (0%)<br>vs.<br>18/21 (86%) controls | 0/1 (0%)<br>vs.<br>14/17 (82%) controls<br><br>35 days post first dose:<br>1/2 (50%)<br>vs.<br>9/13 (69%) controls | |
| Ollila et. al. | Single centre retrospective cohort study<br><br>United States | Haematology Myeloid subset | 160 patients - 6% myeloid | 72 years (65-79) | 86 Male<br>74 Female | BNT162b2<br>mRNA-1273<br>Ad26 | SARS-CoV-2 S IgG Abbott Signal/cutoff ratio ≥1.4 = positive | 56 days post first dose:<br>5/10 (50%) myeloid malignancies | Not reported | Not reported |
| Re et. al. | Multicentre retrospective cohort study<br><br>France | Haematology AML/MDS subset | 102 patients -13% MDS/AML | 76 years (33-93) | 67 Male<br>35 Female | BNT162b2<br>mRNA-1273<br><br>93%/7% | SARS-CoV-2 S IgG Range of commercial kits utilising their threshold | 21-28 days post second dose<br>11/13 (85%) | Not reported | Not reported |

**Supplementary Table 6: Summary of study characteristics and outcomes for patients with acute leukaemia (AML, ALL) and myelodysplastic syndrome (MDS)**

| Study | Type /Location | Study population/ Comparator | Number of participants (analysed) | Age Median | Male/ Female | Vaccine type | Analysis | Seropositivity | Rate of positive neutralising antibody/ cellular response | Adverse events |
|---|---|---|---|---|---|---|---|---|---|---|
| Caocci et. al. | Single centre prospective cohort study<br><br>Italy | MPN | 20 patients<br>-65% MF<br>-30% ET<br>-5% PV | 66 years (48-82) | Not reported | BNT162b2 | SARS-CoV-2 S IgG DiaSorin<br>≥ 15 AU/ml = positive | 42 days post second dose: 13/20 (65%) | Not reported | Not reported |
| Chowdhury et. al. | Single centre retrospective cohort<br><br>United Kingdom | CML and MPN<br><br>Healthcare workers<br>> 60 years old | 59 patients<br><br><br>232 controls | 62 years (IQR 52-73) | 27 Male<br>32 Female | BNT162b2<br>ChAdOx1<br><br>37%<br>63% | SARS-CoV-2 S IgG Abbott<br>≥ 50 AU/mL = positive | ≥ 2 weeks post first dose:<br>34/59 (57%)<br><br>224/232 (97%) | Not reported | Not reported |
| Guglielmelli et. al. | Single centre prospective cohort study<br><br>Italy | MPN<br><br><br><br><br>Healthy controls | 30 patients<br>-43% MF<br>-33% PV<br>-23% ET<br><br>14 controls | Not reported overall | 10 Male<br>20 Female | BNT162b2<br>mRNA-1273<br><br>83%<br>17% | SARS-CoV-2 S/RBD IgG<br>Not specified | 21 to 28 days post first dose:<br>18/30 (60%)<br>vs.<br>14/14 (100%) controls | 21 to 28 days post first dose:<br>13/30 (43%)<br>vs.<br>14/14 (100%) controls | Not reported |
| Harrington et. al. | Single centre prospective cohort study<br><br>United Kingdom | MPN | 16 patients<br>-CML | 45 years (23-74) | 12 Male<br>4 Female | BNT162b2 | SARS-CoV-2<br>S IgG 'in house'<br>1:25 = positive<br><br>SARS-CoV-2 | 21 days post first dose:<br>14/16 (88%) | Neutralising antibody 21 days post first dose:<br>6/16 (38%)<br><br>Cellular:<br>14/15 (80%) | Local adverse events: 8/16 (50%)<br><br>Systemic adverse events: 9/16 (56%) |

| | | | | | | | | | | |
|---|---|---|---|---|---|---|---|---|---|---|
| | | | | | | | neutralising ‘in house’ ID50 = positive<br><br>SARS-CoV-2 T cells ICS (IFN, IL2)<br>3 fold increase = positive | | | |
| Harrington et. al. | Single centre prospective cohort study<br><br>United Kingdom | MPN | 21 patients | 58 years (36-72) | 7 Male<br>21 Female | BNT162b2 | SARS-CoV-2<br>S IgG ‘in house’<br>1:25 = positive<br><br>SARS-CoV-2 neutralising ‘in house’ ID50 = positive<br><br>SARS-CoV-2 T cells ICS (IFN, IL2)<br>3 fold increase = positive | 21 days post first dose: 16/21 (76%) | Neutralising antibody 21 days post first dose: 18/21 (86%)<br><br>Cellular (CD4): 15/20 (75%) | At least 1 adverse event: 12/21 (57%) local<br><br>10/21 (48%) systemic |
| Pimpinelli et. al. | Single centre prospective cohort study | MPN | 42 patients<br>-40% ET<br>-36% PV<br>-24% MF | 72 years (52-82) | 20 Male<br>22 Female | BNT162b2 | SARS-CoV-2 S IgG DiaSorin<br>≥ 15 AU/ml | 21 days post first dose: 23/42 (55%) | Not reported | Not reported |

| | | | | | | | | | | |
|---|---|---|---|---|---|---|---|---|---|---|
| | Italy | | | | | | = positive | 14 days post second dose: 36/42 (86%) | | |
| **Subset of myeloproliferative neoplasm and chronic myeloid leukaemia patients from other haematology studies** | | | | | | | | | | |
| Benda et. al. | Single centre prospective cohort study | Haematology MPN subset<br><br>Solid tumour | 123 patients -28% AML/MDS/ MPN<br><br>136 patients | Not reported in haem | Not reported in haem | BNT162b2 | SARS-CoV-2 S IgG Roche ≥ 0.8 IU/ml = positive | 28 days post second dose: 33/34 (97%) | Not reported | Not reported for haem |
| Greenberger et. al. | Multicentre prospective cohort study<br><br>United States | Haematology CML subset | 1445 patients -2% CML | 68 years (16-110) | 574 Male 871 Female | BNT162b2 mRNA-1273<br><br>45%<br>55% | SARS-CoV-2 S IgG Roche ≥ 0.8 IU/ml = positive | >14 days post second dose: 33/34 (97%) | Not reported | Not reported |
| Herzog Tzarfati et. al. | Single centre prospective cohort study<br><br>Israel | Haematology<br><br>Matched Healthy control | 315 patients -22% MPN -7% CML<br><br>108 controls | 70 years (IQR 61-77) | 223 Male 200 Female | BNT162b2 | SARS-CoV-2 S IgG DiaSorin ≥ 12 AU/ml = positive | 30-60 days post second dose: 57/68 (84%) MPN 20/22 (91%) CML vs. 107/108 (99%) control | Not reported | Not reported |
| Monin et. al. | Multicentre prospective cohort study<br><br>United Kingdom | Haematology MPN subset | 56 patients -68% B cell -9% T cell -18% myeloid, leukaemia -5% others | 73 years (IQR 65-80) | Not extractable for haem | BNT162b2 | SARS-CoV-2 Spike IgG ≥ 70 EC50 = positive<br><br>SARS-CoV-2 specific T | 21 days post first dose: 1/5 (20%) vs. 32/34 (94%) Controls<br><br>35 days post | Cellular response 21 days post first dose: 1/3 (33%) vs. 14/17 (82%) controls | Not extractable for haem |

| | | | | | | | | | | |
|---|---|---|---|---|---|---|---|---|---|---|
| | | Health care workers controls | 54 controls | | | | cells secreting IFN-gamma and/or IL2 >7 cytokine-secreting cells per $10^6$ PBMC = positive | first dose: 1/4 (25%) vs. 18/21 (86%) controls<br><br>35 days post first dose (with second dose): 1/1 (100%) vs. 12/12 (100%) controls | 35 days post first dose: 1/2 (50%) vs. 9/13 (69%) controls<br><br>35 days post first dose (with second dose): 1/1 (100%) vs. 3/3 (100%) controls | |
| Pimpinelli et. al. | Single centre prospective study<br><br>Italy | Haematology MPN subset<br><br>Older age (> 80 years) control group | 50 patients -40% CML -22% ET -22% PV -16% MF<br><br>36 controls | 70 years (28-80) | 26 Male 24 Female | BNT162b2 | SARS-CoV-2 S IgG DiaSorin ≥ 15 AU/ml = positive | 21 days post first dose: 26/50 (52%) MPN vs. 19/36 (53%) controls<br><br>14 days post second dose: 44/50 (88%) MPN vs. 36/36 (100%) controls<br><br>≥ 80 AU/ml 14 days post second dose 42/50 (84%) MPN vs. | Not reported | Reported with different patient numbers |

| | | | | | | | | | | |
|---|---|---|---|---|---|---|---|---|---|---|
| | | | | | | | | 34/36 (97%) control | | |
| Re et. al. | Multicentre retrospective cohort study<br><br>France | Haematology MPN subset | 102 patients<br>-45% lymphoma<br>-22% myeloma<br>-13% MDS/AML<br>-10% CLL<br>-10% MPN | 76 years (33-93) | 67 Male<br>35 Female | BNT162b2<br>mRNA-1273<br><br>93%/7% | SARS-CoV-2 S IgG<br>Range of commercial kits utilising their threshold | 21-28 days post second dose<br>8/10 (80%) | Not reported | Not reported |

**Supplementary Table 7: Summary of study characteristics and outcomes for patients with myeloproliferative neoplasm and chronic myeloid leukaemia**

| Study | Type /Location | Study population/ Comparator | Number of participants (analysed) | Age Median | Male/ Female | Vaccine type | Analysis | Seropositivity | Rate of positive neutralising antibody/ cellular response | Adverse events |
|---|---|---|---|---|---|---|---|---|---|---|
| Avivi et. al. | Single centre Prospective cohort study<br><br>Israel | Myeloma / Healthy volunteers | 171 patients vs. 64 controls | 70 years (range 38-94) | 96 male<br>75 female | BNT162b2 | SARS-CoV-2 S IgG Roche ≥ 0.8 UI/ml = positive | 14-21 days post second dose: 110/147 (75%) active treatment | Not reported | At least one adverse event: 90/161 (53%) vs. 29/53 (55%) controls |
| Bird et. al. | Retrospective cohort study<br><br>United Kingdom/ Europe | Myeloma | 93 patients | 67 years (47-87) | 55 Male<br>38 Female | BNT162b2<br>ChAdOx1<br><br>52%<br>48% | SARS-CoV-2 S IgG Ortho clinical ≥1 signal/cut-off = positive | ≥ 21 days post first dose: 32/66 (48%) on active therapy<br><br>20/27 (74%) not on active therapy | Not reported | Not reported |
| Terpos et. al. | Single centre prospective cohort study<br><br>Greece | Myeloma<br><br><br>Matched Controls | 48 patients<br><br><br>102 controls | 83 years (59-92) | 29 Male<br>19 Female | BNT162b2 | SARS-CoV-2 neutralising Ab Genscript ≥ 30% = positive ≥ 50% = clinically relevant | Not reported | ≥ 50% 22 days post first dose: 4/13(31%) no active therapy | Not reported |
| Terpos et. al. | Single centre prospective cohort study | Myeloma | 276 patients -77% myeloma | 74 years (62-80) | 151 Male<br>125 Female | BNT162b2<br>ChAdOx1 | SARS-CoV-2 neutralising | Not reported for active treatment | Day 50 post first dose: 23/34 (68%) | First dose BNT162b2 71/215 |

| | | | | | | | | | | |
|---|---|---|---|---|---|---|---|---|---|---|
| | Greece | Matched Controls | -14% sMM<br>-9% MGUS<br><br>226 controls | | | 78%/22% | Ab Genscript<br>≥ 30% = positive<br>≥ 50% = clinically relevant | | no active treatment | (33%) local reaction<br><br>28/215 (13%) systemic reaction<br><br>ChAdOx1 20/61 (33%) local reaction<br><br>Second dose BNT162b2 68/215 (32%) local reaction<br><br>45/215 (21%) systemic reaction |
| Van Oekelen et. al. | Single centre prospective and retrospective cohort study<br><br>United States | Myeloma<br><br>Matched control health care workers | 320 patients -260 sampled<br><br>67 controls | 68 years (38-93) | 185 Male<br>135 Female | BNT162b2<br>mRNA-1273<br>unknown<br><br>69%<br>27%<br>4% | SARS-CoV-2 S IgG<br>≥ 5 AU/ml = positive | 51 days post second dose: 43/44 (98%) no active treatment | Not reported | Not reported |
| Benjamini et. al. | Multicentre prospective cohort study<br><br>Israel | CLL patients | 373 patients | 70 years (40-89) | 222 Male<br>151 Female | BNT162b2 | SARS-CoV-2 S IgG DiaSorin<br>≥ 15 AU/ml = positive | 14-21 days pot second dose: 16/120 (13%) active treatment | Not reported | At least 1 adverse event: 151/331 (47%) |

| | | | | | | | | | | |
|---|---|---|---|---|---|---|---|---|---|---|
| | | | | | | | Abbott >50 U/ml = positive ‘in house’ >1.1 = positive | 143/253 (57%) no active treatment | | |
| Del Poeta et. al. | Single centre prospective cohort study<br><br>Italy | CLL patients | 46 patients | Not reported | 29 Male<br>17 Female | BNT162b2 | SARS-CoV-2 S IgG Maglumi ≥1.1 = positive | 14-21 days post second dose:<br>12/29 (41%) on active treatment | Not reported | Not reported |
| Herishanu et. al. | Single centre prospective cohort study<br><br>Israel | CLL patients<br><br>Control -age, sex matched | 167 patients<br><br>52 controls | 71 years (63-76) | 112 Male<br>55 Female | BNT162b2 | SARS-CoV-2 S IgG Roche ≥ 0.8 IU/ml = positive | 14-21 days post second dose:<br>23/58 (55%) treatment naïve patients<br><br>12/75 (16%) Patients on active treatment | Not reported | First dose 52/167 (31%) local reaction<br><br>21/167 (13%) Systemic reaction<br><br>Second dose 56/167 (34%) local reaction<br><br>21/167 (23%) Systemic reaction |
| Roeker et. al. | Single centre retrospective cohort study | CLL patients | 44 patients | 71 years (37-89) | 23 Male<br>21 Female | BNT162b2<br>mRNA-1273 | SARS-CoV-2 S IgG | 21 days post second dose:<br>6/26 (23%) | Not reported | Not reported |

| | | | | | | | | | | |
|---|---|---|---|---|---|---|---|---|---|---|
| | United States | | | | | 57%/43% | DiaSorin ≥ 15 AU/ml = positive | not on active treatment<br><br>17/18 (94%) treatment naïve | | |
| Tadmor et. al. | Multicentre prospective observation study<br><br>Israel | CLL patients | 84 patients | Not reported overall | 53 Male*<br>29 Female | BNT162b2 | SARS-CoV-2 S IgG Abbott ≥ 50 U/ml = positive<br><br>SARS-CoV-2 RBD IgG >1.1 = positive | Day 22 post second dose:<br>6/21 (29%) on active treatment<br><br>43/63 (68%) not on active treatment | Not reported | Not reported |
| Lim et. al. | Multicentre prospective cohort study<br><br>United Kingdom | Lymphoma<br><br><br>Healthy control | 129 patients recruited<br>119 analysed<br>-66% indolent B-NHL<br>-29% aggressive B-NHL<br>-10% HL<br>-3% other<br><br>150 control | 69 years (IQR 57-74) | 81 Male<br>48 Female | BNT162b2<br>ChAdOx1 | SARS-CoV-2 S IgG Meso Scale Discovery >0.55 BAU/ml = positive<br><br>RBD IgG >0.73 BAU/ml = positive | 14 days post first dose:<br>9/31 (28%) on active treatment<br><br>23/28 (82%) not on active treatment<br><br>14-28 days post second dose:<br>13/33 (39%) on active treatment<br><br>48/53 (91%) not on active | Not reported | Not reported |

| | | | | | | | | | | |
|---|---|---|---|---|---|---|---|---|---|---|
| | | | | | | | | treatment | | |
| Agha et. al. | Single centre retrospective cohort study<br><br>United States | Haematology | 67 patients | 71 years (IQR 65-77) | 35 Male<br>32 Female | BNT162b2<br>mRNA-1273<br><br>51%/42%<br>7% unknown | SARS-CoV-2 S IgG Beckman Coulter ≥1.00 = positive | 21 days post second dose:<br>15/29 (52%) Active treatment<br><br>21/38 (55%) not on active treatment | Not reported | Not reported |
| Greenberger et. al. | Multicentre prospective cohort study<br><br>United States | Haematology | 1445 patients<br>-45% CLL<br>-25% NHL<br>-5% HL<br>-15% myeloma<br>-4% acute leukaemia<br>-2% CML<br>-2% MPN<br>-2% others | 68 years (16-110) | 574 Male<br>871 Female | BNT162b2<br>mRNA-1273<br><br>45%<br>55% | SARS-CoV-2 S IgG Roche ≥ 0.8 IU/ml = positive | >14 days post second dose:<br>669/747 (90%) not on active treatment | Not reported | Not reported |
| Harrington et. al. | Single centre prospective cohort study<br><br>United Kingdom | MPN | 21 patients | 58 years (36-72) | 7 Male<br>21 Female | BNT162b2 | SARS-CoV-2 S IgG 1:25 = positive<br><br>SARS-CoV-2 neutralising ID50 = positive<br><br>SARS-CoV-2 T cells | 21 days post first dose:<br>4/7 (57%) active treatment<br><br>12/14 (86%) not on active treatment | Not reported | At least 1 adverse event:<br>12/21 (57%) local<br><br>10/21 (48%) systemic |

| | | | | | | | | | | |
|---|---|---|---|---|---|---|---|---|---|---|
| | | | | | | | ICS (IFN, IL2)<br>3 fold increase = positive | | | |
| Herzog Tzarfati et. al. | Single centre prospective cohort study<br><br>Israel | Haematology<br><br>Matched Healthy control | 315 patients<br>-22% MPN<br>-17% Myeloma<br>-16% aggressive NHL<br>-13% indolent NHL<br>-11% CLL<br>-7% CML<br>-5% HL<br>-5% Acute leukemia<br>-5% MDS<br><br>108 controls | 70 years (IQR 61-77) | 223 Male<br>200 Female | BNT162b2 | SARS-CoV-2 S IgG DiaSorin<br>≥ 12 AU/ml = positive | 30-60 days post second dose:<br>130/151 (86%) not on active treatment | Not reported | Not reported |
| Jurgens et. al. | Single centre prospective cohort study<br><br>United States | Haematology<br><br>Control health care workers | 67 patients<br>-31% CLL<br>-63% NHL<br>-6% HL<br><br>35 controls | 71 years (24-90) | 36 Male<br>31 Female | BNT162b2<br>mRNA-1273<br><br>46%<br>54% | SARS-CoV-2 S IgG 'in house'<br>OD450 ≥ 3 = positive | 21 days post second dose:<br>9/29 (31%) on active treatment | Not reported | Not reported |
| Ollila et. al. | Single centre retrospective cohort study<br><br>United States | Haematology | 160 patients<br>-36% aggressive lymphoma<br>-21% | 72 years (65-79) | 86 Male<br>74 Female | BNT162b2<br>mRNA-1273<br>Ad26 | SARS-CoV-2 S IgG Abbott Signal/cuto | 56 days post first dose:<br>10/15 (67%) in patients under | Not reported | Not reported |

| | | | | | | | | | | |
|---|---|---|---|---|---|---|---|---|---|---|
| | | | indolent lymphoma<br>-15% plasma cell<br>-12% CLL<br>- 9% other lymphoma<br>- 6% myeloid | | | | ff ratio ≥1.4 = positive | observation/no treatment<br><br>31/63 (49%) patients in remission post treatment | | |
| Re et. al. | Multicentre retrospective cohort study<br><br>France | Haematology | 102 patients<br>-45% lymphoma<br>-22% myeloma<br>-13% MDS/AML<br>-10% CLL<br>-10% MPN | 76 years (33-93) | 67 Male<br>35 Female | BNT162b2<br>mRNA-1273<br><br>93%/7% | SARS-CoV-2 S IgG<br>Range of commercial kits utilising their threshold | 21-28 days post second dose<br>31/47 (66%) no active treatment | Not reported | Not reported |

**Supplementary Table 8: Summary of study characteristics and outcomes for subgroup analysis active treatment vs. no active treatment**

| Study | Type /Location | Study population/ Comparator | Number of participants (analysed) | Age Median | Male/ Female | Vaccine type | Analysis | Seropositivity | Rate of positive neutralising antibody/ cellular response | Adverse events |
|---|---|---|---|---|---|---|---|---|---|---|
| Benjamini et. al. | Multicentre prospective cohort study<br><br>Israel | CLL patients | 373 patients | 70 years (40-89) | 222 Male<br>151 Female | BNT162b2 | SARS-CoV-2 S IgG<br>DiaSorin ≥ 15 AU/ml = positive<br>Abbott >50 U/ml = positive<br>'in house' >1.1 = positive | 14-21 days pot second dose:<br>2/39 (5%) CD20 <12 months<br>32/92 (35%) CD20 >12 months | Not reported | At least 1 adverse event: 151/331 (47%) |
| Herishanu et. al. | Single centre prospective cohort study<br><br>Israel | CLL patients<br><br>Control -age, sex matched | 167 patients<br><br>52 controls | 71 years (63-76) | 112 Male<br>55 Female | BNT162b2 | SARS-CoV-2 S IgG<br>Roche ≥ 0.8 IU/ml = positive | 14-21 days post second dose:<br>0/22 (0%) treatment with CD20 < 12 months<br><br>25/55 (46%) if CD20 ≥ 12 months | Not reported | First dose<br>52/167 (31%) local reaction<br>21/167 (13%) Systemic reaction<br><br>Second dose<br>56/167 (34%) local reaction<br>21/167 (23%) Systemic reaction |

| | | | | | | | | | | |
|---|---|---|---|---|---|---|---|---|---|---|
| Roeker et. al. | Single centre retrospective cohort study<br><br>United States | CLL patients | 44 patients | 71 years (37-89) | 23 Male<br>21 Female | BNT162b2<br>mRNA-1273<br><br>57%/43% | SARS-CoV-2 S IgG DiaSorin ≥ 15 AU/ml = positive | 21 days post second dose: 2/14 (14%) if CD20 < 12 months | Not reported | Not reported |
| Tadmor et. al. | Multicentre prospective observation study<br><br>Israel | CLL patients | 84 patients | Not reported overall | 53 Male*<br>29 Female | BNT162b2 | SARS-CoV-2 S IgG Abbott ≥ 50 U/ml = positive<br><br>SARS-CoV-2 RBD IgG >1.1 = positive | Day 22 post second dose: 18/22 (82%) if CD20 > 12 months<br><br>0/22 (0%) CD20 <12 months | Not reported | Not reported |
| Gurion et. al. | Multicentre prospective cohort study<br><br>Israel | Lymphoma | 162 patients<br>-88% NHL<br>-12% HL | 65 years (52-73) | 89 Male<br>73 Female | BNT162b2 | SARS-CoV-2 S IgG Abbott ≥ 50 IU/ml = positive | 28 days post second dose: 25/66 (38%) CD20 < 12 months<br><br>17/21 (81%) if CD20 > 12 months | Not reported | Not reported |
| Easdale et. al. | Single centre retrospective cohort study<br><br>United Kingdom | Allogeneic HCT<br>>3 months | 55 patients | 50 years (18-73) | 34 Male<br>21 Female | BNT162b2<br>ChAdOx1<br><br>38%/62% | SARS-CoV-2 S IgG Ortho clinical ≥1 signal/cut-off = | 42 days post first dose: 2/10 (20%) if CD20 < 12 months | Not reported | Not reported |

| | | | | | | | | | | |
|---|---|---|---|---|---|---|---|---|---|---|
| | | | | | | | positive | | | |
| Agha et. al. | Single centre retrospective cohort study<br><br>United States | Haematology | 67 patients | 71 years (IQR 65-77) | 35 Male<br>32 Female | BNT162b2<br>mRNA-1273<br><br>51%<br>42%<br>7% unknown | SARS-CoV-2 S IgG Beckman Coulter ≥1.00 = positive | 21 days post second dose: 2/9 (22%) CD20 < 12 months | Not reported | Not reported |
| Greenberger et. al. | Multicentre prospective cohort study<br><br>United States | Haematology | 1445 patients<br>-45% CLL<br>-25% NHL<br>-5% HL<br>-15% myeloma<br>-4% acute leukaemia<br>-2% CML<br>-2% MPN<br>-2% others | 68 years (16-110) | 574 Male<br>871 Female | BNT162b2<br>mRNA-1273<br><br>45%<br>55% | SARS-CoV-2 S IgG Roche ≥ 0.8 IU/ml = positive | >14 days post second dose: 97/263 (37%) on CD20 | Not reported | Not reported |
| Herzog Tzarfati et. al. | Single centre prospective cohort study<br><br>Israel | Haematology | 315 patients<br>-22% MPN<br>-17% Myeloma<br>-16% aggressive NHL<br>-13% indolent NHL<br>-11% CLL<br>-7% CML<br>-5% HL<br>-5% Acute leukemia<br>-5% MDS | 70 years (IQR 61-77) | 223 Male<br>200 Female | BNT162b2 | SARS-CoV-2 S IgG DiaSorin ≥ 12 AU/ml = positive | 30-60 days post second dose: 0/28 (0%) on CD20 | Not reported | Not reported |

| | | | | | | | | | | |
|---|---|---|---|---|---|---|---|---|---|---|
| | | Matched Healthy control | 108 controls | | | | | | | |
| Jurgens et. al. | Single centre prospective cohort study<br><br>United States | Haematology<br><br><br><br><br>Control health care workers | 67 patients<br>-31% CLL<br>-63% NHL<br>-6% HL<br><br>35 controls | 71 years (24-90) | 36 Male<br>31 Female | BNT162b2<br>mRNA-1273<br><br>46%/54% | SARS-CoV-2 S IgG<br>'in house'<br>OD450 ≥ 3 = positive | 21 days post second dose:<br>0/8 (0%) on CD20 | Not reported | Not reported |
| Re et. al. | Multicentre retrospective cohort study<br><br>France | Haematology | 102 patients<br>-45% lymphoma<br>-22% myeloma<br>-13% MDS/AML<br>-10% CLL<br>-10% MPN | 76 years (33-93) | 67 Male<br>35 Female | BNT162b2<br>mRNA-1273<br><br>93%/7% | SARS-CoV-2 S IgG<br>Range of commercial kits utilising their threshold | 21-28 days post second dose<br>1/17 (6%) if on CD20<br><br>14/22 (64%) if CD20 ≥ 12 months | Not reported | Not reported |
| Thakkar et. al. | Single centre prospective and retrospective cohort study<br><br><br>United States | Haematology<br><br><br><br><br><br><br><br>Solid tumours<br><br><br>Healthy control | 66 patients<br>-39% lymphoid<br>-27% myeloid<br>-33% plasma cell<br><br>134 patients<br><br><br>26 controls | Not reported for haem | Not reported for haem | BNT162b2<br>mRNA-1273<br>Ad26 | SARS-CoV-2 S IgG<br>Abbott<br>≥ 50 AU/ml = positive | 29 days post completion (2 doses for mRNA, 1 dose for Ad26):<br>4/5 (80%) CD20 < 12 months<br><br>12/18 (67%) CD20 > 12 months | Not reported | Not reported for haem |

**Supplementary Table 9: Summary of study characteristics and outcomes for subgroup analysis CD20 therapy less than 12 months vs. CD20 therapy 12 or more months**

| Study | Type /Location | Study population/ Comparator | Number of participants (analysed) | Age Median | Male/ Female | Vaccine type | Analysis | Seropositivity | Rate of positive neutralising antibody/ cellular response | Adverse events |
|---|---|---|---|---|---|---|---|---|---|---|
| Benjamini et. al. | Multicentre prospective cohort study<br><br>Israel | CLL patients | 373 patients | 70 years (40-89) | 222 Male<br>151 Female | BNT162b2 | SARS-CoV-2 S IgG DiaSorin ≥ 15 AU/ml = positive Abbott >50 U/ml = positive 'in house' >1.1 = positive | 14-21 days pot second dose:<br>14/79 (18%) BTKi<br>2/34 (6%) venetoclax<br>7/49 (14%) venetoclax plus CD20 | Not reported | At least 1 adverse event: 151/331 (47%) |
| Del Poeta et. al. | Single centre prospective cohort study<br><br>Italy | CLL patients | 46 patients | Not reported | 29 Male<br>17 Female | BNT162b2 | SARS-CoV-2 S IgG Maglumi ≥1.1 = positive | 14-21 days post second dose:<br>8/21 (38%) BTKi<br>2/8 (25%) Venetoclax | Not reported | Not reported |
| Herishanu et. al. | Single centre prospective cohort study<br><br>Israel | CLL patients<br><br>Control -age, sex matched | 167 patients<br><br>52 controls | 71 years (63-76) | 112 Male<br>55 Female | BNT162b2 | SARS-CoV-2 S IgG Roche ≥ 0.8 IU/ml = positive | 14-21 days post second dose:<br>8/50 (16%) BTKi<br><br>3/20 (14%) venetoclax +/- CD20<br><br>2/5 (40%) venetoclax | Not reported | First dose 52/167 (31%) local reaction<br><br>21/167 (13%) Systemic reaction<br><br>Second dose |

| | | | | | | | | | | |
|---|---|---|---|---|---|---|---|---|---|---|
| | | | | | | | | | | 56/167 (34%) local reaction<br><br>21/167 (23%) Systemic reaction |
| Parry et. al. | Single centre prospective cohort study<br><br>United Kingdom | CLL patients<br><br><br>Healthy age matched controls | 299 patients<br><br><br>93 controls | 69 years (IQR 63-74) | 159 Male<br>140 Female | BNT162b2<br>ChAxOd1<br><br>52%<br>48% | SARS-CoV-2 S IgG<br>Roche<br>≥ 0.8 IU/ml = positive | 43 days post first dose:<br>Serum<br>2/18 (11%) BTKi | Not reported | Not reported |
| Roeker et. al. | Single centre retrospective cohort study<br><br>United States | CLL patients | 44 patients | 71 years (37-89) | 23 Male<br>21 Female | BNT162b2<br>mRNA-1273<br><br>57%/43% | SARS-CoV-2 S IgG<br>DiaSorin<br>≥ 15 AU/ml = positive | 21 days post second dose:<br>3/14 (21%) BTKi<br>0/7 (0%) Venetoclax | Not reported | Not reported |
| Tadmor et. al. | Multicentre prospective observation study<br><br>Israel | CLL patients | 84 patients | Not reported overall | 53 Male<br>29 Female | BNT162b2 | SARS-CoV-2 S IgG<br>Abbott<br>≥ 50 U/ml = positive<br><br>SARS-CoV-2 RBD IgG >1.1 = positive | Day 22 post second dose:<br>4/11 (36%) BTKi<br><br>1/6 (17%) venetoclax | Not reported | Not reported |

| | | | | | | | | | | |
|---|---|---|---|---|---|---|---|---|---|---|
| Greenberg er et. al. | Multicentre prospective cohort study<br><br>United States | Haematology | 1445 patients<br>-45% CLL<br>-25% NHL<br>-5% HL<br>-15% myeloma<br>-4% acute leukaemia<br>-2% CML<br>-2% MPN<br>-2% others | 68 years (16-110) | 574 Male<br>871 Female | BNT162b2<br>mRNA-1273<br><br>45%<br>55% | SARS-CoV-2 S IgG Roche<br>≥ 0.8 IU/ml = positive | >14 days post second dose:<br>136/286 (48%) BTKi<br><br>11/28 (39%) venetoclax | Not reported | Not reported |
| Herzog Tzarfati et. al. | Single centre prospective cohort study<br><br>Israel | Haematology<br><br><br><br><br><br><br><br><br><br><br>Matched Healthy control | 315 patients<br>-22% MPN<br>-17% Myeloma<br>-16% aggressive NHL<br>-13% indolent NHL<br>-11% CLL<br>-7% CML<br>-5% HL<br>-5% Acute leukemia<br>-5% MDS<br><br><br>108 controls | 70 years (IQR 61-77) | 223 Male<br>200 Female | BNT162b2 | SARS-CoV-2 S IgG DiaSorin<br>≥ 12 AU/ml = positive | 30-60 days post second dose:<br>2/5 (60%) BTKi<br>1/4 (25%) venetoclax | Not reported | Not reported |
| Jurgens et. al. | Single centre prospective cohort study<br><br>United States | Haematology | 67 patients<br>-31% CLL<br>-63% NHL<br>-6% HL | 71 years (24-90) | 36 Male<br>31 Female | BNT162b2<br>mRNA-1273<br><br>46%<br>54% | SARS-CoV-2 S IgG 'in house' OD450 ≥ 3 | 21 days post second dose:<br>6/10 (60%) BTKi | Not reported | Not reported |

| | | | | | | | | | | |
|---|---|---|---|---|---|---|---|---|---|---|
| | | Control health care workers | 35 controls | | | | = positive | | | |
| Re et. al. | Multicentre retrospective cohort study<br><br>France | Haematology | 102 patients<br>-45% lymphoma<br>-22% myeloma<br>-13% MDS/AML<br>-10% CLL<br>-10% MPN | 76 years (33-93) | 67 Male<br>35 Female | BNT162b2<br>mRNA-1273<br><br>93%<br>7% | SARS-CoV-2 S IgG<br>Range of commercial kits utilising their threshold | 21-28 days post second dose<br>27/36 (75%) if on targeted therapy | Not reported | Not reported |

**Supplementary Table 10: Summary of study characteristics and outcomes for subgroup analysis targeted therapy vs. no targeted therapy**

| Study | Type /Location | Study population/ Comparator | Number of participants (analysed) | Age Median | Male/ Female | Vaccine type | Analysis | Seropositivity | Rate of positive neutralising antibody/ cellular response | Adverse events |
|---|---|---|---|---|---|---|---|---|---|---|
| Bird et. al. | Single centre retrospective cohort study<br><br>United Kingdom/ Europe | Myeloma | 93 patients | 67 years (47-87) | 55 Male<br>38 Female | BNT162b2<br>ChAdOx1<br><br>52%/48% | SARS-CoV-2 S IgG Ortho clinical ≥1 signal/cut-off = positive | ≥ 21 days post first dose:<br>6/8 (75%) HCT </= 12 months<br><br>37/69 (54%) HCT > 12 months | Not reported | Not reported |
| Dhakal et. al. | Single centre retrospective cohort study<br><br>United States | Autologous<br>Allogeneic HCT<br>CAR-T | 130 patients<br>-45 autoHCT<br>-71 alloHCT<br>-14 CAR-T | 65 years (45-75)<br>64 years (25-77) | Not reported | BNT162b2<br>mRNA-1273<br>Ad26 | SARS-CoV-2 S IgG EUROIMMUN ≥1.1 signal/cut-off = positive | 14 days post completion of vaccination:<br><br>AutoHCT<br>11/15 (73%) < 12 months<br>16/30 (53%) ≥ 12 months<br><br>AlloHCT<br>11/19 (58%) < 12 months<br>38/52 (73%) ≥ 12 months | Not reported | Not reported |
| Thakkar et. al. | Single centre prospective and retrospective cohort study | Haematology | 66 patients<br>-39% lymphoid<br>-27% myeloid | Not reported for haem | Not reported for haem | BNT162b2<br>mRNA-1273<br>Ad26 | SARS-CoV-2 S IgG Abbott ≥ 50 AU/ml | 29 days post completion (2 doses for mRNA, 1 dose for | Not reported | Not reported for haem |

| | | | | | | | | | | |
|---|---|---|---|---|---|---|---|---|---|---|
| | United States | Solid tumours<br><br>Healthy control | -33% plasma cell<br><br>134 patients<br><br>26 controls | | | | = positive | Ad26):<br>2/3 (67%) if HCT < 12 months<br><br>17/23 (74%) HCT > 12 months | | |
| Van Oekelen et. al. | Single centre prospective and retrospective cohort study<br><br>United States | Myeloma<br><br>Matched control health care workers | 320 patients -260 sampled<br><br>67 controls | 68 years (38-93) | 185 Male<br>135 Female | BNT162b2<br>mRNA-1273<br>unknown<br><br>69%<br>27%<br>4% | SARS-CoV-2 S IgG<br>≥ 5 AU/ml = positive | 51 days post second dose:<br>9/9 (100%) if HCT < 12 months | Not reported | Not reported |

**Supplementary Table 11: Summary of study characteristics and outcomes for subgroup analysis haematopoietic stem cell transplant within 12 months vs. 12 or more months**

| Study | Type /Location | Study population/ Comparator | Number of participants (analysed) | Age Median | Male/ Female | Vaccine type | Analysis | Seropositivity | Rate of positive neutralising antibody/ cellular response | Adverse events |
|---|---|---|---|---|---|---|---|---|---|---|
| Bird et. al. | Retrospective cohort study<br><br>United Kingdom/ Europe | Myeloma | 93 patients | 67 years (47-87) | 55 Male<br>38 Female | BNT162b2<br>ChAdOx1<br><br>52%<br>48% | SARS-CoV-2<br>S IgG<br>Ortho clinical<br>≥1 signal/cut-off = positive | ≥ 21 days post first dose:<br>26/48 (54%) BNT162b2<br><br>26/45 (58%) ChAdOx1 | Not reported | Not reported |
| Chowdhury et. al. | Single centre retrospective cohort<br><br>United Kingdom | CML and MPN<br><br>Healthcare workers > 60 years old | 59 patients<br><br>232 controls | 62 years (IQR 52-73) | 27 Male<br>32 Female | BNT162b2<br>ChAdOx1<br><br>37%<br>63% | SARS-CoV-2 S IgG Abbott<br>≥ 50 AU/mL = positive | ≥ 2 weeks post first dose:<br>12/22 (55%) BNT162b2<br><br>22/37 (59%) ChAdOx1 | Not reported | Not reported |
| Lim et. al. | Multicentre prospective cohort study<br><br>United Kingdom | Lymphoma Subset off treatment | 129 patients recruited<br>119 analysed<br>-66% indolent B-NHL<br>-29% aggressive B-NHL<br>-10% HL<br>-3% other<br><br>150 control | 69 years (IQR 57-74) | 81 Male<br>48 Female | BNT162b2<br>ChAdOx1 | SARS-CoV-2 S IgG Meso Scale Discovery<br>>0.55 BAU/ml = positive<br><br>RBD IgG<br>>0.73 BAU/ml = positive | 14 days post first dose:<br>14/18 (78%) BNT162b2<br><br>9/10 (90%) ChAdOx1<br><br>14-28 days post second dose:<br>31/35 (89%) | Not reported | Not reported |

| | | | | | | | | | | |
|---|---|---|---|---|---|---|---|---|---|---|
| | | Healthy control | | | | | | BNT162b2<br><br>17/18 (94%) ChAdOx1 | | |
| Van Oekelen et. al. | Single centre prospective and retrospective cohort study<br><br>United States | Myeloma<br><br>Matched control health care workers | 320 patients -260 sampled<br><br>67 controls | 68 years (38-93) | 185 Male 135 Female | BNT162b2 mRNA-1273 unknown<br><br>69%<br>27%<br>4% | SARS-CoV-2 S IgG ≥ 5 AU/ml = positive | 51 days post second dose: 144/147 (98%) BNT162b2<br><br>68/76 (89%) mRNA-1273 | Not reported | Not reported |

**Supplementary Table 12: Summary of study characteristics and outcomes for subgroup analysis vaccine type (BNT162b2 vs others)**

| | **All studies (n=44)** | | | **Good and fair quality studies (n=23)** | | |
|---|---|---|---|---|---|---|
| | Single arm studies<br><br>Pooled response rate (95% CI) | Intervention arm of comparator studies<br><br>Pooled response rate (95% CI) | Intervention vs. control cohort<br><br>Odds ratio (95% CI) Heterogeneity *p*-value | Single arm studies<br><br>Pooled response rate (95% CI) | Intervention arm of comparator studies<br><br>Pooled response rate (95% CI) | Intervention vs. control cohort<br><br>Odds ratio (95% CI) Heterogeneity *p*-value |
| Following second dose | 0.61 (0.54-0.69)<br><br>$I^2$ = 92%, $p$<0.01 | 0.67 (0.58-0.76)<br><br>$I^2$ = 93%, $p$<0.01 | OR 0.05 (0.02-0.15)<br>$p$<0.01<br>$I^2$ = 88%, $p$<0.01 | 0.63 (0.52-0.75)<br><br>$I^2$ = 90%, $p$<0.01 | 0.66 (0.51-0.81)<br><br>$I^2$ = 94%, $p$<0.01 | OR 0.08 (0.01-0.58)<br>$p$=0.02<br>$I^2$ = 90%, $p$<0.01 |
| Following first dose | 0.51 (0.38-0.64)<br><br>$I^2$ = 92%, $p$<0.01 | 0.37 (0.23-0.51)<br><br>$I^2$ = 90%, $p$<0.01 | OR 0.10 (0.04-0.29)<br>$p$<0.01<br>$I^2$ = 86%, $p$<0.01 | 0.53 (0.33-0.72)<br><br>$I^2$ = 93%, $p$<0.01 | 0.30 (0.09-0.51)<br><br>$I^2$ = 90%, $p$<0.01 | OR 0.31 (0.13-0.78)<br>$p$=0.03<br>$I^2$ = 62%, $p$=0.05 |

**Supplementary Table 13: Summary of seropositivity rates for patients with haematological malignancy following 2 and 1 dose of COVID-19 vaccine by study quality (sensitivity analysis)**

### Additional information: Search strategy

Database: Ovid MEDLINE(R) ALL <1946 to August 31, 2021>
Search Strategy:
--------------------------------------------------------------------------------
1 exp hematologic neoplasms/ or exp leukemia/ or exp lymphoma/ or exp multiple myeloma/ or exp myeloproliferative disorders/ or exp myelodysplastic-myeloproliferative diseases/ or exp Stem Cell Transplantation/ or exp bone marrow transplantation/ (554418)
2 ((h?ematologic* or hematopoietic or blood or bone marrow) adj3 (cancer* or neoplasm* or malignan* or carcinoma*)).mp. (56690)
3 (leuk?emia* or lymphoma* or myeloma* or hodgkin* or myelodysplastic or myeloproliferative or polycythemia vera or stem cell transplant* or bone marrow transplant* or chimeric antigen receptor therap* or CAR-T).mp. (723006)
4 1 or 2 or 3 (758330)
5 exp COVID-19 vaccines/ or ((exp vaccines/ or exp immunization/) and (COVID-19/ or SARS-CoV-2/)) (6448)
6 ((covid* or SARS-CoV-2 or coronavirus or BNT162b2 or ChAdOx1 or AZD 1222, or mRNA-1273 or Ad26* or Ad5* or NVX-CoV2373 or pfizer or astrazeneca or astra-zeneca or oxford or novavax or moderna or johnson) adj2 (vaccin* or immuniz* or immunis*)).mp. (9371)
7 5 or 6 (11023)
8 4 and 7 (191)
9 limit 8 to english language (185)
10 limit 9 to yr="2020 -Current" (185)

***************************

Database: Embase <1974 to 2021 August 31>
Search Strategy:
--------------------------------------------------------------------------------
1 exp hematologic malignancy/ or exp leukemia/ or exp lymphoma/ or exp myeloma/ or exp myeloproliferative disorder/ or exp myelodysplastic syndrome/ or exp Stem Cell Transplantation/ or exp bone marrow transplantation/ (867821)
2 ((h?ematologic* or hematopoietic or blood or bone marrow) adj3 (cancer* or neoplasm* or malignan* or carcinoma*)).mp. (95692)
3 (leuk?emia* or lymphoma* or myeloma* or hodgkin* or myelodysplastic or myeloproliferative or polycythemia vera or stem cell transplant* or bone marrow transplant* or chimeric antigen receptor therap* or CAR-T).mp. (1032174)
4 1 or 2 or 3 (1097575)
5 exp SARS-CoV-2 vaccine/ or ((exp vaccine/ or exp immunization/) and (exp coronavirus disease 2019/ or exp Severe acute respiratory syndrome coronavirus 2/)) (12263)
6 ((covid* or SARS-CoV-2 or coronavirus or BNT162b2 or ChAdOx1 or AZD 1222, or mRNA-1273 or Ad26* or Ad5* or NVX-CoV2373 or pfizer or astrazeneca or astra-zeneca or oxford or novavax or moderna or johnson) adj2 (vaccin* or immuniz* or immunis*)).mp. (9360)
7 5 or 6 (14554)
8 4 and 7 (339)
9 limit 8 to english language (333)
10 limit 9 to yr="2020 -Current" (329)

**************************

Cochrane CENTRAL

Search Name: Covid-19 vaccine haem SR
Last Saved: 31/08/2021 13:15:41

ID Search
#1 (h?ematologic* or hematopoietic or blood or bone marrow) near/3 (cancer* or neoplasm* or malignan* or carcinoma*)
#2 (leuk?emia* or lymphoma* or myeloma* or hodgkin* or myelodysplastic or myeloproliferative or polycythemia vera or stem cell transplant* or bone marrow transplant* or chimeric antigen receptor therap* or CAR-T)
#3 #1 or #2
#4 (covid* or SARS-CoV-2 or coronavirus or BNT162b2 or ChAdOx1 or AZD 1222, or mRNA-1273 or Ad26* or Ad5* or NVX-CoV2373 or pfizer or astrazeneca or astra-zeneca or oxford or novavax or moderna or johnson) near/2 (vaccin* or immuniz* or immunis*)
#5 #3 and #4 with Cochrane Library publication date in The last 2 years

**Additional information: Abbreviations**

CLL: chronic lymphocytic leukaemia; NHL: non hodgkins lymphoma; HL: hodgkins lymphoma; AML: acute myeloid leukaemia; MDS: myelodysplastic syndrome; MM: myeloma; MGUS: monoclonal gammopathy of unknown significance; WG: Waldenstrom's magroglobulinaemia; HCT: haematopoietic stem cell transplantation; CAR-T: chimeric antigen receptor T cell; MPN: myeloproliferative neoplasm; CML: chronic myeloid leukaemia; ET: Essential thrombocytosis; PV: polycythaemia vera; MF: myelofibrosis; AU: arbitrary unit

BNT162b2: Tozinameran (Pfizer); mRNA1273: Spikevax (Moderna); ChAdOx1: Vaxzevria (AstraZeneca); Ad26: Janssen
